# Epstein-Barr Virus Selects for Stem Cell Lineages in Breast Cancer and Normal Tissues

**DOI:** 10.1101/2025.08.25.25334378

**Authors:** Bernard Friedenson

## Abstract

Epstein-Barr virus (EBV) causes persistent damage to breast cancer genomes, so viral particles need not be continually present to cause malignancy. Because basal type stem cells are present in breast cancer but missing in normal breast, the hypothesis was tested that EBV creates or selects for breast cancer stem cells. Bioinformatic comparisons identified methylation of cis-regulators of gene expression in breast cancer chromosomes that closely matched EBV-associated cancers. On certain chromosomes, most of these methylated regulators governed stem cell related differentiation pathways. The shared methylation sites were found in breast cancers typical of the basal layer, where breast stem cells are thought to originate. A few of the same shared breast cancer methylation marks were detected in non-malignant keratinocytes with a cured EBV infection, suggesting EBV epigenetic changes are difficult to reverse. Control experiments with methylation sites on chromosome 17 in (non-EBV) skin cell carcinoma did not correspond to abnormal methylation in breast cancer. Moreover, randomly generated epigenetic sites showed negligible overlap with EBV cancers, so the observed matches are unlikely in non-EBV cancers and unlikely due to chance. Additionally, breast cancer genomes had variable damage to immune responses typical of EBV infection, so immune variation helps explain why not everyone gets EBV cancer. Coupled with other breast cancer safeguards compromised by EBV, epigenetic changes in stem cell related genes that match EBV cancers make the breast more susceptible to cancer triggers.

## Introduction

Saliva, blood, breast milk **[1]**, and semen transmit Epstein-Barr virus (EBV). Transmission is so effective that nearly everyone acquires EBV infection at some point in their lives [2]. The infection provides no known benefit to its host and sometimes causes nasopharyngeal cancer (NPC) and Burkitt’s lymphoma (BL). NPC and endemic BL do not occur without EBV infection, so they are unequivocal examples of EBV-cancers. EBV attaches to human chromosomes as circular episomes which replicate along with host DNA, but the episomes can be lost during replication in NPC and BL cells [3, 4].

In previous work, NPC and BL were used as cancer genome models to determine whether there are EBV-related changes in breast cancer genomes. Breast cancer genomes were found to have critical chromosome damage resembling EBV-induced damage [5, 6]. Genomic comparisons found that thousands of breakpoints and other lesions in breast cancer closely matched EBV-associated cancers. Many of these chromosome lesions are pathogenic and would accelerate breast cancer even if the infection clears or EBV episomes are lost. This persistent damage explains why EBV particles need not be consistently detected in breast cancer [5, 6].

Breast cancer [7], NPC and BL [8–10] all arise from cancer stem cells – the rare cells within a tumor that can self-renew and initiate new tumors [11, 12]. This common origin suggests the hypothesis that breast cancer arises because EBV disrupts normal breast stem cell lineages. If this is true, lesions in breast cancer that interfere with stem cell lineages might match EBV cancers [5]. The virus becomes most successful if the cells it infects cannot be correctly replaced so EBV selects for cells that have damaged or lost the ability to restore themselves correctly.

Stem cells were previously believed to become malignant through the same mechanisms as somatic cells. Stem cells were thought to be more vulnerable than somatic cells because of their plasticity, self-renewal, and longevity. However, stem cells are either absent or vanishingly rare in healthy breast tissue [13]. Single cell transcriptomics of over 700,000 normal breast cells found no clear evidence of a stem cell population with a unique transcription profile[13]. No proliferating cells were detected in basal cells or cell states with stem cell markers, challenging the concept that a basal stem cell fuels epithelial homeostasis. In contrast, the ability to proliferate enabled the identification of progenitors in the luminal compartment. One of these populations of progenitor cells aligned with hormone dependent proliferation [13]. Results from transcriptomics of normal breast cells create a paradox when compared to transcriptomics of breast cancers. In contrast to the absence of canonical stem cells in the basal layer of the normal breast, basal layers in breast cancers show strong evidence for stem cells or their descendants. For instance, the cancer subtype containing basal single cells had the presumptive progenitor cell of origin for basal breast cancer [14]. Stem cells were also present in cells in the cancer microenvironment. One state of cancer associated fibroblasts in the cancer stroma “had features of mesenchymal stem cells with high expression of stem cell markers (ALDH1A1, KLF4, and LEPR).” [14]. Perivascular-like cells in the stroma also expressed markers related to stem cells [14].

EBV infection might resolve this paradox if it provides a mechanism to produce or select for malignant stem cells. EBV infection actively reshapes host cell genomes[5] and epigenomes to select for cells that facilitate viral replication [15]. A precondition for this reshaping is a mechanism of immune escape that releases control of EBV infection. This requirement contributes to explaining why not everyone with EBV infection gets cancer. In NPC and BL, massive changes in host cell methylation [16, 17] then occur as EBV changes accessibility to chromatin, reprogramming cancer epigenomes [18, 19]. Methylation patterns are strong predictors of gene expression levels [20, 21] and unambiguous correlations have been established between cis-methylation sites and genes expressed in breast cancers [22]. Reprogramming can affect stem cell differentiation pathways and the maintenance and regeneration of tissue damage [23]. If this scenario applies to breast cancer, then some breast cancer methylation sites should correspond to those in chromosomes from EBV-associated cancers. The matching sites should be related to genes that regulate stem cell behavior at steps prior to their terminal differentiation. These changes may cause long-lasting damage to breast cancer genomes so that viral particles need not be continually present to cause malignancy.

The research presented below tested these hypotheses. DNA methylation was observed in regions of breast cancer chromosomes that regulate stem cell differentiation, function, and microenvironment. This methylation does not exist in normal cells, but was found in identical or virtually identical positions in prototype EBV-cancers. The agreement in completely unrelated data was so strong and so consistent with established effects of EBV that it supports the hypothesis that EBV selects for epigenetic modifications in stem cells. This selection prevents replacement of abnormal cells and facilitates viral replication. Genes affected include HOX genes and those encoding for morphogens. Differentiation pathways become blocked or deregulated. At least one large subset of breast cancers in the cohort had diverse immune escape mechanisms [24], so serious that they would allow EBV infection. Malignant transformation also creates a homing microenvironment that facilitates growth and multiplication of cancer stem cell seeds in multiple tissues. This milieu provides nutrition, cytokines, and protection from immune responses [25].

## Results

### Methylation sites in breast cancer that control critical genes align with methylation sites in EBV cancers

The minimum distance between abnormal methylation at breast cancer gene control sites and the EBV cancers NPC and BL was calculated for all 22 human autosomes. The most frequent agreement was within 10 base pairs and included the two EBV cancers (Fig. 1, blue and orange bars in 16 of 22 chromosomes). The positions of the shared methyl groups were sometimes exactly the same.

**Fig. 1.**
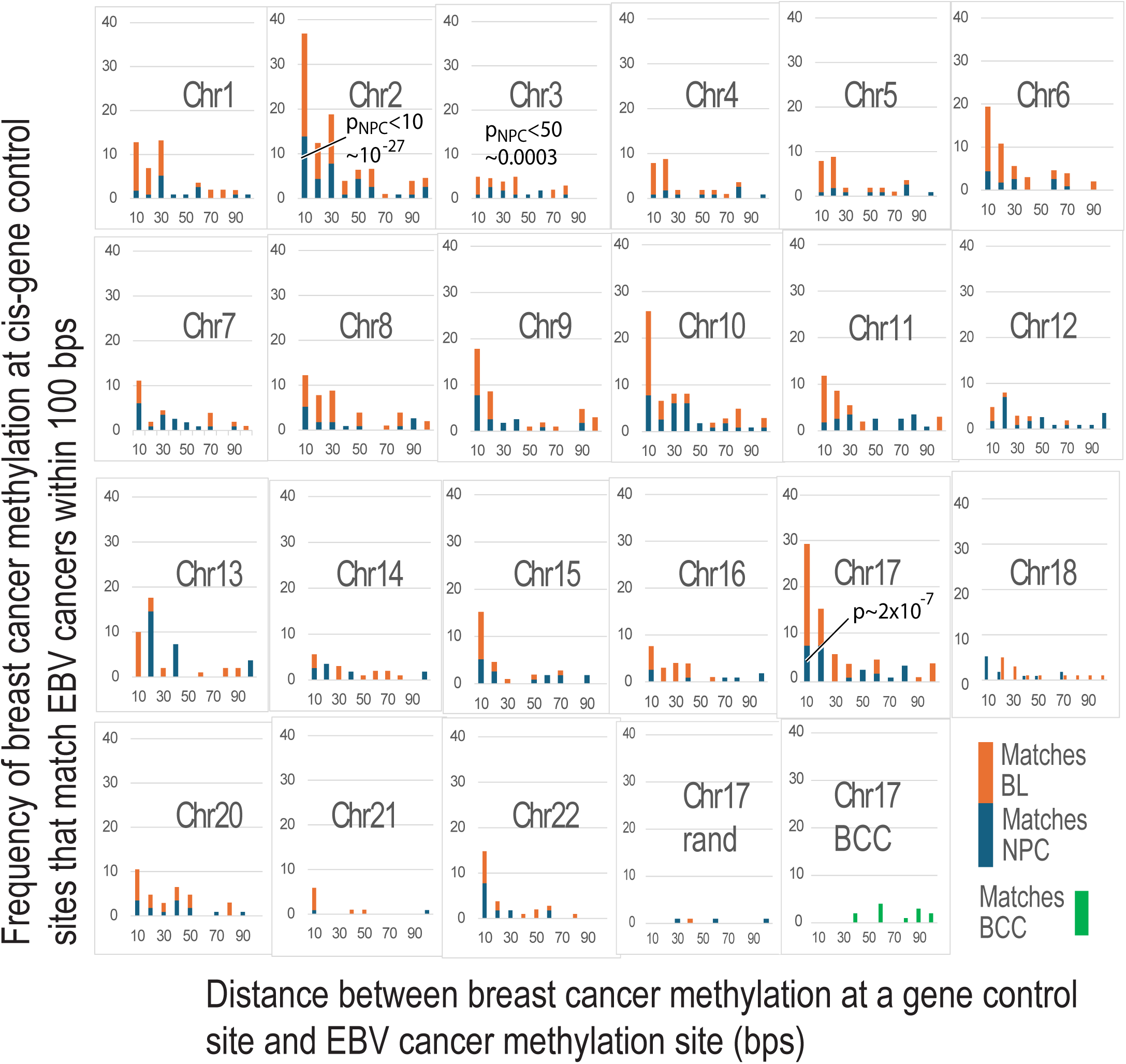
Minimum distance between abnormal methylation sites in breast cancer and EBV cancers. On most chromosomes, abnormal breast cancer methylation sites are within 10 base pairs of an abnormal methylation site in the EBV cancers NPC and BL. Comparison of abnormal methylation sites on chromosome 17 to a random distribution (lower right) in a sample trial shows almost no matches, while other trials were blank. The non-EBV cancer BCC (green bars) also does not resemble the matches on chr17. Multiple comparisons were used to find values that agreed and P values for the comparisons with NPC methylation sites are shown for chromosomes 2, 3, and 17.

Other calculations also showed that chance occurrence was unlikely and supported similarity of abnormal methylation distributions in breast and EBV cancers. For example, comparing 2319 abnormal methylation positions in NPC to 789 breast cancer positions on chromosome 2(242,696,762 bases) found 15 positions that agreed within 10 base pairs while 0.158 matches would be expected by chance. The probability of observing k = 15 or more matches with λ = 0.158 under a Poisson distribution model gives p ≈ 1.3 × 10⁻²⁷. This value is so low it strongly suggests the observed agreement is not random. The same calculation using 9 matches that were found <10 bases apart on chromosome 17 gave p ≈ 2×10^-7^. Chromosome 3 had fewer matches than either chromosome 2 or 17 but 5 matches were <50 bases apart and still statistically significant (p ≈ 0.0003). In addition Mann-Whitney comparisons of the entire distributions of abnormal methylation in breast cancer vs. NPC on seven chromosomes (2, 7, 9,10-12, and 20) give high P-values, which indicate that there is no significant difference between the distributions of abnormal methylation groups. On chromosomes 9, 10, and 11, the confidence interval for the median difference included zero, further suggesting that the medians were not significantly different. Even when the methylation sites on a chromosome near each other were sparse, Mann-Whitney comparisons still could not exclude the possibility that abnormally methylated DNA base distributions were the same.

Ten trials using random data rather than breast cancer data produced blank or nearly blank results (e.g. Fig. 1, lower right). Another control compared these abnormal methylation sites to 13,766 methylation sites on chromosome 17 [26] in basal cell carcinoma of the skin (BCC). The skin cancers had not been corrected by subtracting methylation in normal cells, so much larger numbers of methylation sites had to be compared to breast cancer. Nonetheless matching sites were much rarer and weaker in comparison to breast cancer vs NPC or BL (Fig. 1, lower right). Mann-Whitney testing of abnormal methylation sites in breast cancer and BCC gave statistical evidence that the distributions were different (two-sided p<0.0001 with a 60.7% probability that a randomly selected value from BCC is greaten than one from breast cancer).

These results show that breast cancer chromosomes have frequent abnormal methylation at virtually the same positions as cancers known to be caused by EBV. The sharing of methylation loci between breast and EBV cancers suggests the abnormal methyl groups are essential to produce all three cancers. To characterize this effect, the genes in breast cancers established as controlled by cis-methylation [22] at loci matching EBV cancers were next determined.

### Stem cell related gene controls predominate at sites methylated in the breast cancer positions that match EBV cancers

The next analyses asked whether there were identifiable functions that were preferentially targeted in the breast cancer methylated gene regulators that matched EBV-cancers. Chromosome 17 was the first chromosome evaluated because it encodes the *BRCA1* and *ERBB2 (HER2)* genes, which both have significant roles in breast cancer. Chromosome 17 in the breast cancer cohort [22] had 405 gene entries with abnormal methylation at their regulatory sites and 85% of sequences that regulated these genes were <=200 bp in length.

Methylation sites on chromosome 17 in breast cancer [22], NPC [18], and BL [19] that were not present in normal cells were compared in detail. The first hint that abnormal methylation affected the same functions in the three cancers was found in regulatory regions of *SOX9* (Fig. 2). *SOX9* counterbalances the effect of the core stem cell gene *SOX2*, which regulates pluripotency. In cancer, *SOX9* and *SOX2* are epigenetic regulators that together determine cancer cell plasticity and metastatic progression [27, 28]. In addition, an intricate signaling channel network governs the self-renewal and lineage determination of mammary stem cells. *SOX9* is in this network and controlled by *NOTCH* signaling [29]. *KRT19*, a control point for *NOTCH* signaling, is methylated at positions within 3 bases in breast cancer and BL. Breast cancer and EBV-related cancers had another link to stem cells through shared methylated clusters that affected homeobox genes *HOXB2* and *HOXB4*. *HOX* genes produce a code that contributes to driving tissue stem cells to differentiate into the correct lineages in order to maintain and repair their respective tissues and organs [30]. *HOX* genes are often regulated by morphogen gradients (e.g. Wnt, BMP, FGF, RA, Shh) and are inhibited by epigenetic marks until functional differentiation begins at gastrulation. The marks are removed on differentiation and *HOX* genes become active to guide development of various body regions [31]. As shown in the first panel in Fig. 2, *HOXB2* and *HOXB4* control sites in breast cancers with abnormal methylation at sites that closely correspond to EBV cancers.

**Fig 2.**
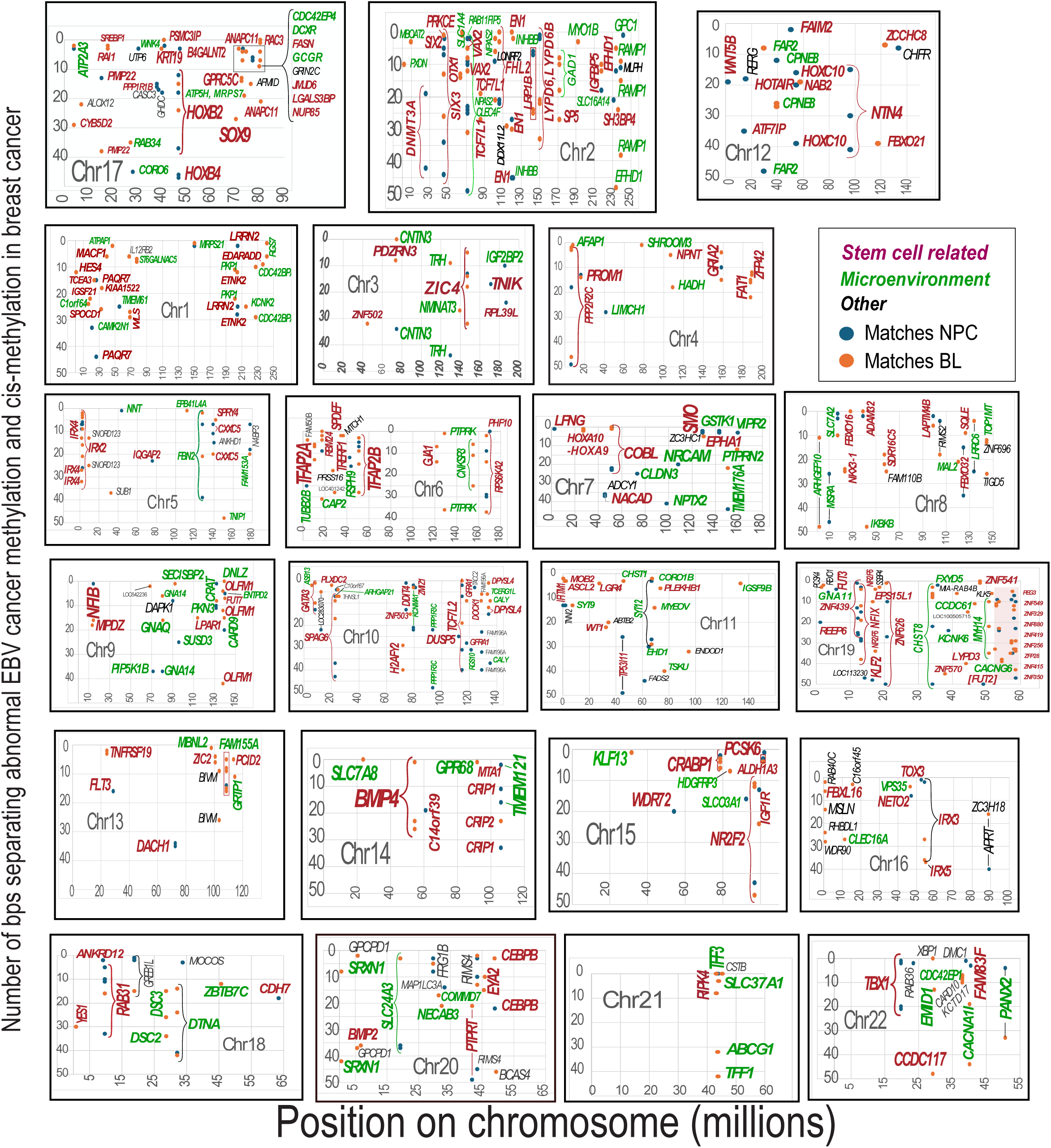
Methylation of DNA controlling breast cancer stem cell genes is different from normal and closely matches the EBV cancers NPC and BL (blue and orange dots, respectively. See the legend in the second row). Genes related to stem cells (red text) or those controlling the environment (green text) are the predominant genes in breast cancer that are close to positions with abnormal methylation in EBV associated cancers. Genes that affect other functions are indicated in black. Distances on the vertical axis represent positions of abnormal cis-methylation in breast cancer and EBV cancers that agree within <50 bp. Many positions are identical or almost identical for all three types of cancers (top of each panel). The three chromosomes in the top row are described in the text. The red shaded area in chromosome 19 for zinc finger proteins infers matches, but the data is incomplete.

As a control, the number of CpG islands in NPC DNA sequences that matched breast cancer sequences were calculated as 100 times the numbers of CpG islands in DNA divided by the length of DNA. All the comparisons at any window size involved dense numbers of CpG islands in the differentially methylated DNA, ranging from 24 to 7.4, that did not depend on the length of DNA. *HOXB2* gave a value of 16.5 and *NUP85* gave a low value of 7.4. Other regions on chromosome 17 gave values as low as 1.31. The lengths of matching DNA sequences were as low as 42 base pairs (*HOXB2*). These results show that relatively short CpG-rich DNA segments were selected for unusual methylation in known EBV mediated cancers and some of these segments closely matched breast cancer cis gene control regions.

Finding *SOX9, KRT19,* and *HOX* genes with abnormal methylation at about the same positions in both breast and EBV cancers raised the possibility that EBV induces methylation that interferes with breast stem cell differentiation. Data in Fig. 1 hint at this possibility. Although young and old stem cells are similar, the numbers of stem cell and their potential cell fates decline with age. The number of abnormal methyl groups in children with BL was almost always higher than in those that matched adults with NPC. Based on this observation, subsequent analyses pursued the possibility that EBV had also targeted stem cells on the other autosomal chromosomes in breast cancers.

### Breast cancer stem cell genes are selected at methylation sites that match EBV-cancers

Essentially identical abnormal methylation positions in breast cancer and EBV cancers extended to other chromosomes with preferential targeting of stem cell related genes or transcription factors that influence cell lineage. Supplementary Table S1 compiles the functions of about 350 of these genes. As shown by the red text in Fig. 2, breast cancer gene promoters and enhancer regions are within 50 bps of methylated positions in EBV cancers. These matching methylated gene regulators affect morphogen responsive *HOX* genes and genes with other associations to stem cells on all twenty-two non-sex chromosomes.

For chromosome 17 (Fig. 3 and supplementary Table S1), some relationship to stem cells was found for about 66% of abnormal methylated positions in breast cancer cis gene control positions corresponding to EBV cancers. Across all 22 chromosomes, 48% of these genes were related to stem cells or to pathways essential for stem cell differentiation. Multiple chromosomes showed methylation controls for genes that disrupt cell fate decisions, self renewal, and tissue homeostasis. A few examples below illustrate these effects.

**Fig. 3.**
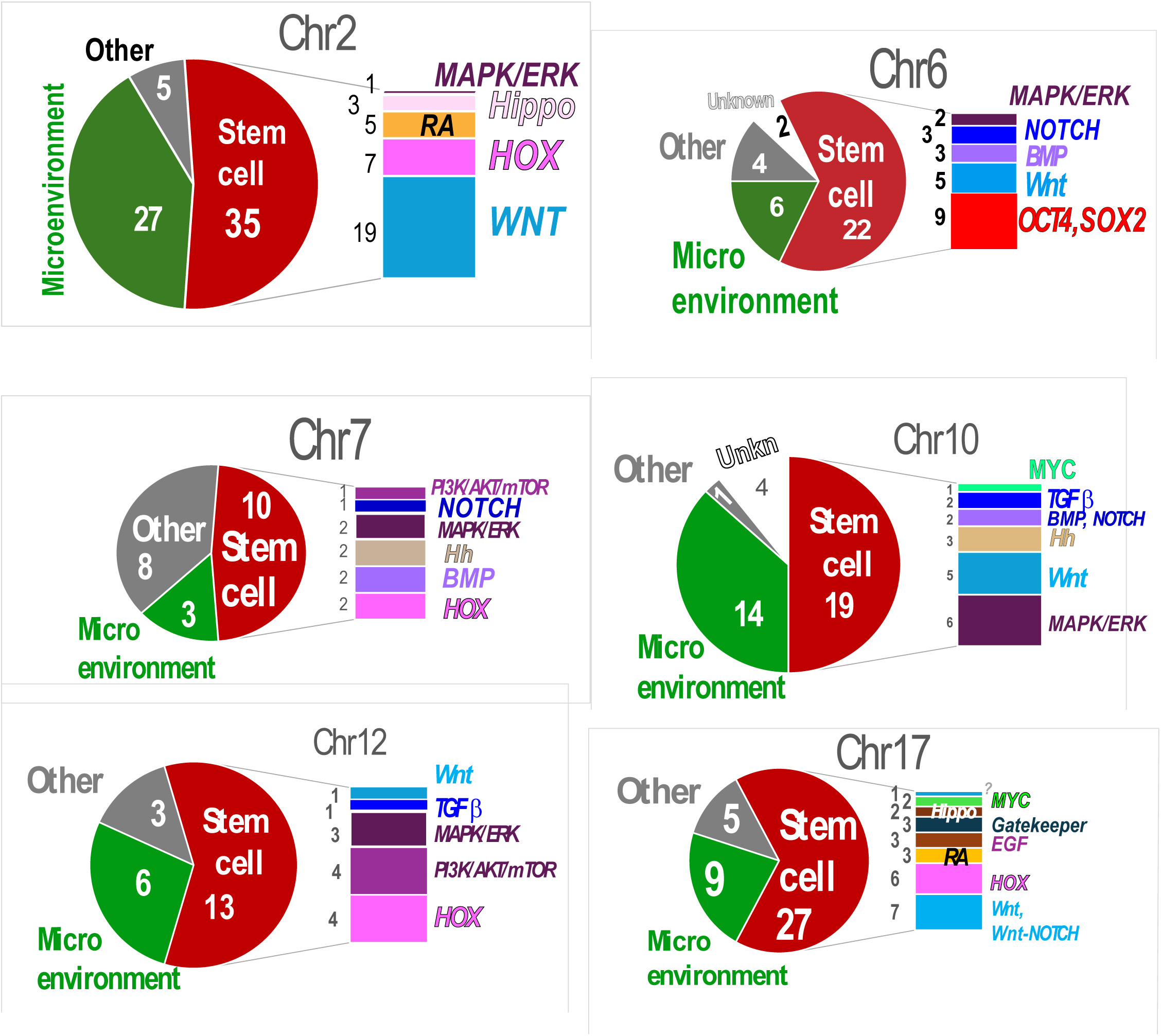
On some chromosomes, most breast cancer gene control regions with abnormal cis-methylation corresponding to EBV cancers affect stem cell differentiation pathways and morphogenesis. Occurrences of shared abnormal methylation loci impacting stem cells, microenvironment or other properties are indicated on the pie charts. Differentiation pathways most frequently affected by the breast cancer cis-methylation sites that match EBV cancer are shown in bar charts to the right of the pie charts. The number of times a given stem cell differentiation pathway is affected by shared abnormal methylation is given at the left of the bar chart. Abnormal breast cancer methylation on chromosome 6 that matches EBV cancers directly affects the pluripotency genes OCT4 and SOX2.

Chromosome 2 genes with altered controls in breast cancers matching EBV cancers had strong relationships to stem cells (Figs 2,3 and Table S1). Genes that shared methylation loci included *SIX2*, *SIX3*, *VAX2*, and *EN1*. While *HOX* genes are master regulators of stem cell destinations and identity, *SIX, VAX,* and *EN* genes participate in finer regulation in specific tissues and regions during development. All these genes nevertheless activate or suppress responses to morphogen gradients during development [GeneCards] and to maintain tissue homeostasis [32]. A different control exists on chromosome 12 where the *HOTAIR* gene contributes to regulating *HOX* expression. *HOTAIR* is 2200 bases of antisense non-coding RNA that inhibits *HOXD* cluster transcription. *HOTAIR* binds to specific chromatin modification complexes to regulate chromatin. Interactions with *HOTAIR*/*PRC* control *HOXD* gene transcriptional activity on chromosome 2.

Supplementary Table S1. gives 167 instances of abnormal methylation that mark stem cell related functions or differentiation pathways with significant specificity in the breast, nasopharynx, or hematopoietic system. These examples suggest that pluripotent cells in breast cancer may have been infected and modified by EBV.

### Differentiation pathways affected by similarities in abnormal methylation

To further investigate the effects of EBV on stem cell pluripotency, the frequency was estimated of differentiation pathways with abnormal EBV-related methylation sites. Six chromosomes were tested based on numerous close (<10 bp) matches to EBV cancers in positions of abnormal methylation and the known distribution of *HOX* genes. As shown in Fig. 3, morphogen associated pathways had abnormal methylation, but the sets of pathways differed on different chromosomes. Only the predominant morphogen connected pathways are indicated, but abnormal methylation usually had links to multiple additional pathways in different contexts. On chromosome 6, the stem cell genes *OCT4* and *SOX2* were the most frequent targets. These two gene cooperate and are essential to reprogram somatic cells to maintain and regain pluripotent stem cells. Their expression is under tight control, and their deregulation can collapse the entire pluripotency network [33, 34]. In addition, *HOX* gene controls were abnormally methylated on four chromosomes, where they were frequent targets. Other genes connected to Wnt and other stem cell differentiation pathways are also predominant targets. These results show that abnormal methylation in breast cancers at sites closely related to EBV can have a profound effect on stem cell pluripotency and the ability to regenerate tissue damage.

### Effect of methylation on gene expression at breast cancer gene control sites related to EBV

In the METABRIC cohort, correlations exist between cis-specific (same chromosomes) methylation and expression “at hundreds of promoters and over a thousand distal elements” [22]. Effects of these cis-methylations near those in EBV cancers were tested. Expression levels were available for 20,603 genes from 1980 breast cancer patients in the METABRIC cohort [35]. As shown in Fig. 4, abnormal methylation on established cis gene control regions and related to EBV cancers more often caused over-expression of stem cell related genes, but under expression also occurred frequently. The most highly over-expressed and under-expressed genes (>50 instances) were labeled in Fig. 4 and they include specific links to cancer development in the context of stem cells. Table 1 groups these abnormally expressed genes according to whether they are generally pluripotency factors, morphogens, homeobox genes, or other differentiation regulators in stem cells.

**Fig. 4.**
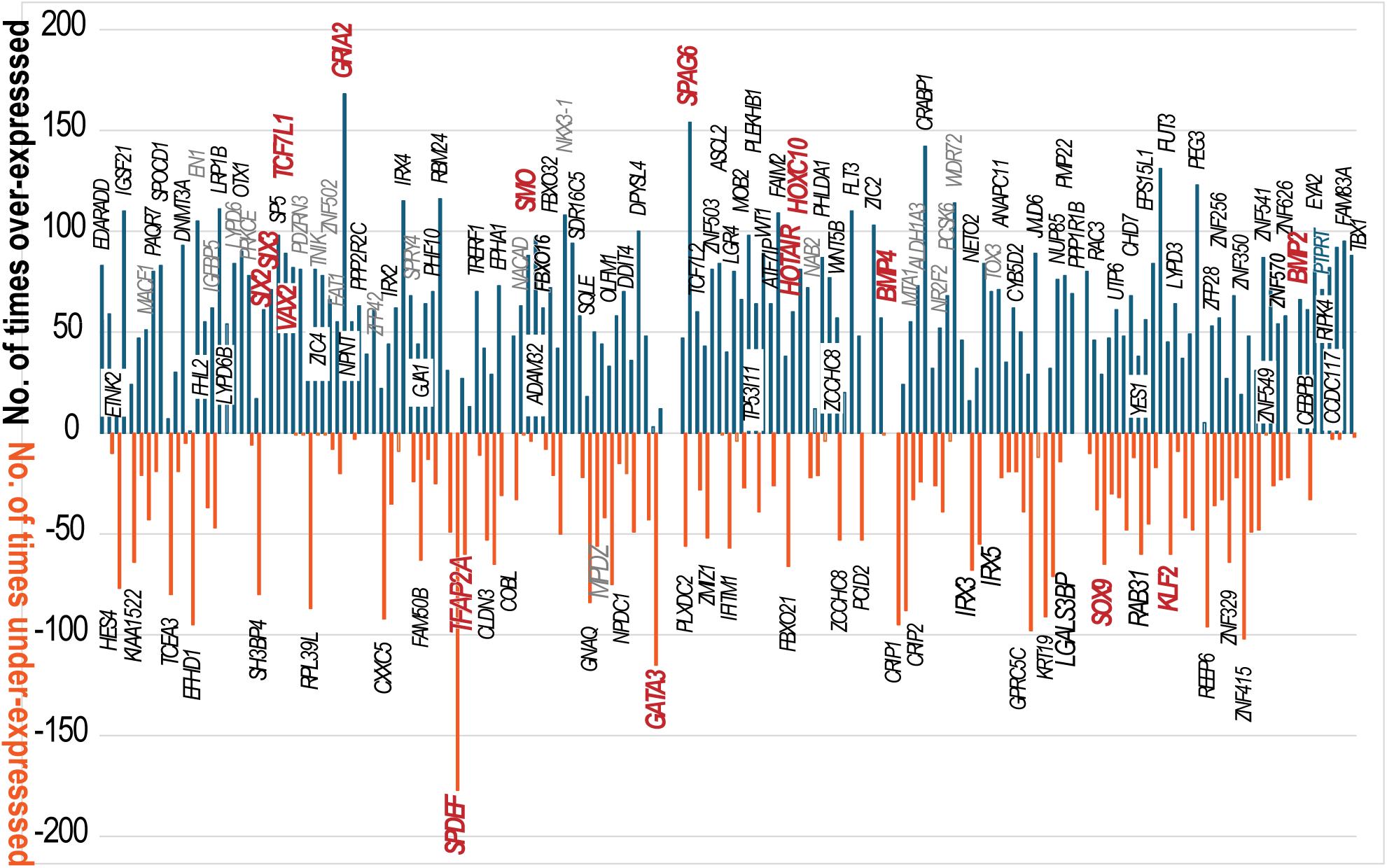
Genes with abnormal methylation in breast cancers matching EBV cancers (+-50 base pairs) are over or under expressed. The number of times these genes are over or under expressed in the METABRIC cohort is shown on the vertical axis. Genes over-or under-expressed more than 50 times are labeled on the 22 autosomes. Genes mentioned in the text as examples are highlighted in red.

**Table 1.** Effects on stem cells by over– and under-expressed genes in breast cancers. Abnormal methylation of these gene control sites is closely related to abnormal methylation sites in EBV cancers

| Gene | Chr | General description of function | Expression Status in breast cancers | Context and reference |
| --- | --- | --- | --- | --- |
| **BMP2** | 20 | Morphogen | Overexpressed | Drives differentiation |
| **BMP4** | 14 | Morphogen | Overexpressed | Drives differentiation |
| **CRABP1** | 15 | Morphogen | Overexpressed | Powerful regulator of stem cell fate via retinoic acid signaling, diminishes neural and embryonic stem cell expansion [72]. |
| **GRIA2** | 4 | Differentiation regulator | Overexpressed | Neurodevelopment gene. Steers progenitors toward specific cell fates after an injury [73] |
| **HOTAIR** | 12 | Homeobox gene regulator | Overexpressed | Epigenetic regulator Causes H3K27 methylation; increases breast cancer invasiveness |
| **HOXC10** | 12 | Homeobox gene | Overexpressed | Activates *ERK*, *AKT*, *p65*, and EMT-related genes |
| **KLF2** | 19 | Pluripotency regulator [38]. | Overexpressed | Increases expression of stem cell markers like OCT4 and NANOG, and suppresses differentiation markers to maintain stemness and self-renewal [74] |
| **SIX2** | 2 | Homeobox gene | Overexpressed | Enhances expression of stem cell markers *SOX2* and *Nanog* [75, 76] Activates Wnt/β-catenin and MAPK pathways; |
| **SIX3** | 2 | Homeobox gene | Overexpressed | Modulates Wnt signaling; enhances proliferation and migration in gastric cancer [77] |
| **SMO** | 7 | Morphogen-responsive gene | Overexpressed | Guides differentiation into specific identities |
| **SOX9** | 17 | Pluripotency regulator | Overexpressed | Antagonizes and opposes pluripotency factor *SOX2* [37] |
| **SPAG6** | 10 | Differentiation regulator | Overexpressed | *SPAG6* is a cilia-related gene. Its overexpression biases stem cells toward specific lineages, and inhibits proliferation of neural progenitors [78]. |
| ***SPDEF*** | 6 | Differentiation regulator (epithelial) | Under-expressed (most significantly in Fig. 4) | Crucial in epithelial cell differentiation. *SPDEF* under-expression during androgen deprivation therapy increased TGF-beta signaling activity to promote epithelial mesenchymal transition (which is strongly linked to stem cell properties) [79] |
| ***TCF7L1*** | 2 | Pluripotency regulator vs. self renewal | Overexpressed | Silences Wnt targets; maintains pluripotent state and prevents differentiation [39] |
| ***TCF7L2*** | 2 | Differentiation regulator | Overexpressed | In many types of tumors, *TCF7L2* promotes tumor invasion and metastasis, often via epithelial mesenchymal transition [80]. Can either activate or repress Wnt signaling |
| ***TFAP2A*** | 6 | Pluripotency exit regulator | Under-expressed | Methylation locks cells in self-renewal; disrupts placental lineage commitment [42]. |
| ***TFAP2B*** | 6 | Differentiation regulator | Under-expressed | Determines differentiation outcome and cell fate in an embryonic melanocyte stem cell [81]. Suppresses Wnt/β-catenin pathways in certain cancers [82]. Not as well studied as *TFAP2A* |
| ***VAX2*** | 2 | Homeobox gene | Overexpressed | Translates positional info from *HOX* genes |

Some over-expressed genes (highlighted in red in Fig. 4) can contribute to creating breast cancer stem cells, either leaving cells in a progenitor-like state so that they do not mature into specialized types or reprogramming them to ectopic expression [36]. The genes labeled as pluripotency regulators include *SOX9* and *KLF2*. *SOX9* opposes the pluripotency factor *SOX2* and pushes cells to differentiate [37], while *KLF2* maintains self renewal and stemness [38]. Overexpressed *TCFL1* in stem-like cancer cells keeps them in a pluripotent state, unable to differentiate by silencing Wnt targets [39].

Other highlighted over-expressed genes include morphogens (e.g. *BMP2* on chromosome 20*, BMP4* on chromosome 14, and morphogen responsive genes (e.g. *SMO* on chromosome 7) that direct differentiation into specific cellular identities. On chromosome 2, *SIX2* [40], *SIX3* and *VAX2* are over-expressed homeobox genes that help translate positional information from *HOX* genes into development.

On chromosome 6, abnormal methylation causes under-expression of *TFAP2A.* Normally TFAP2A combines with OCT4 and SOX2 proteins to initiate exit from pluripotency [41], so methylation of *TFAP2A* regulatory elements locks cells into a self-renewing state. In a different context, *TFAP2A* also cooperates with *GATA2/3* and *TFAP2C* in the “Tetra” network. This network represses pluripotency genes like *OCT4* and couples this repression with initiating differentiation toward the placental support system. Under-expressed *TFAP2A* disrupts this network, preventing proper lineage commitment [42]. *TFAP2A* binds to epigenetically inactive placental genes when their differentiation begins. Without *TFAP2A* contributing to activating these genes, they remain silent, and differentiation stops.

*HOXC10* (located on chromosome 12) expression is normally low in most differentiated tissues, but its over-expression can activate *ERK, AKT,* and *p65* oncogenic pathways and epithelial mesenchymal transition-related genes [43]. *HOTAIR* is encoded within the same *HOXC* gene cluster on chromosome12. *HOTAIR* over-expression causes histone H3K27 methylation, which increases breast cancer invasiveness [44].

Further examples are also included and summarized in Table 1. Over-expressed and under-expressed genes at EBV methylation sites on any of multiple chromosomes can reprogram transcriptional networks, affecting hundreds of downstream genes involved in differentiation and malignancy.

### Control Experiment showing both ER-Negative and ER-positive breast Cancer Cells have methylation in positions similar to EBV-cancers

Many estrogen receptor (ER)-negative breast cancer cells are believed to originate from the basal layer of mammary epithelium. Basal-like breast cancers typically lack expression of ER, PR (progesterone receptor), and HER2, and are often referred to as triple-negative breast cancers. They are highly heterogeneous and notoriously hard to treat. These basal cells are located in the outer layer of the mammary ducts and are distinct from luminal cells, which typically give rise to ER-positive cancers. The basal layer contains stem and progenitor cells as candidates for the origin of aggressive subtypes of basal-like breast cancer.

Conventional stem cells are absent from the basal layer of normal adult breast tissue [13], yet breast cancers exhibit stem-like cells that are the presumptive origin of basal breast cancers [14]. Both normal and cancer datasets agree that there are stem like cells in the luminal layer. If EBV selects or produces basal stem cell phenotypes, then ER negative and ER positive breast cancers should have abnormal methylation in stem cell genes that matches methylation sites in EBV cancers. As shown in Fig. 5, ER-negative breast cancers have from 10 to 52% of methylation near methylation sites in EBV cancers on various chromosomes. Thus EBV methylation sites may select or produce cancer stem-like cells that originate breast cancer in both basal type and luminal layers.

**Fig. 5.**
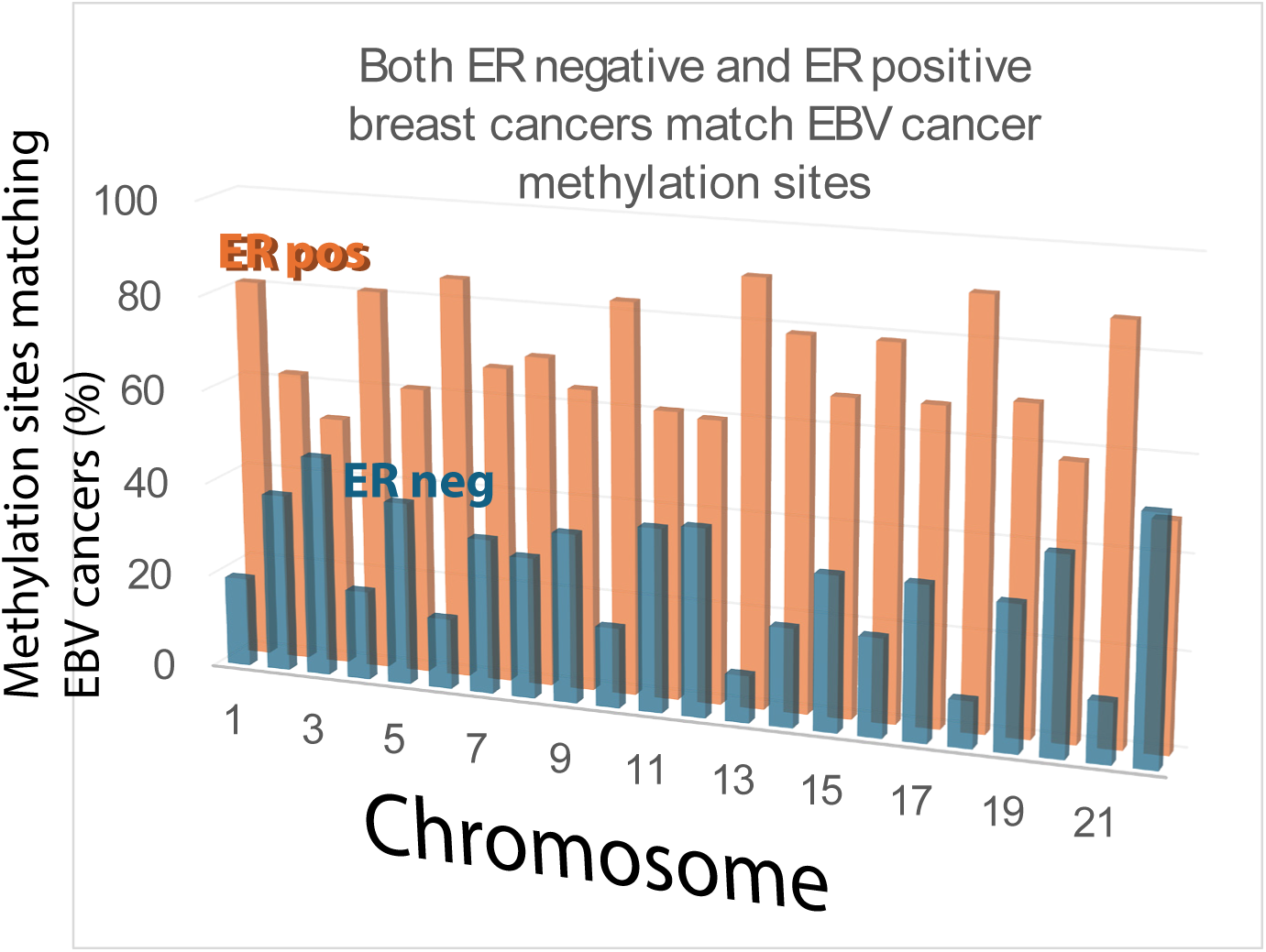
Both ER negative (typically basal type) and ER positive (typically luminal type) breast cancers have abnormal cis-methylation in sites that closely match cancers associated with EBV. This is true for all 22 autosomes and the matching sites are pre-dominantly stem cell related genes.

### Non-malignant cells from people who have recovered from an EBV infection exhibit abnormal DNA chromosome methylation in a few of the same loci as breast cancer cells

To what extent do the results thus far apply to non-malignant cells with an active or a resolved past EBV infection? Are gene control sites methylated by viral infection in non-malignant cells related to those in EBV cancer? DNA methylation in EBV-infected oral keratinocytes differs from uninfected oral keratinocytes and produces DNA scars after the virus is cleared [45]. To determine if these infection related methylation scars were related to breast cancers, the distributions of methyl groups in breast cancer cells were compared to cured, but previously infected oral keratinocytes.

Linear regression analysis revealed a correlation between epigenetic marks in these cell types (r=0.93, r2=0.87, p<0.0001) with a power for 5% significance of 99.99%. Kolmogorov-Smirnov tests (p=0.89) and Spearman’s rank correlation (p=0.88, CI=0.72-0.95) gave consistent results. These results show that methylation in previously infected non-malignant cells correlates with methylation in breast cancer cells. Again these results are unlikely due to pure chance. For example, chromosome 22 has 299 methylation sites in oral keratinocytes. Methylation that controls the RAB36 gene is 7 bases from one of 136 methylation sites in breast cancer (Fig. 6) and other genes that align are closer. The number of sites that agree expected by pure chance is 15X136×299/51324926=0.0119 which gives a p value of 0.012, assuming a Poisson distribution. The p values for chromosome 22 show that pure chance cannot explain the relatively rare agreement between non-malignant keratinocytes with a past EBV infection and breast cancers.

**Fig. 6.**
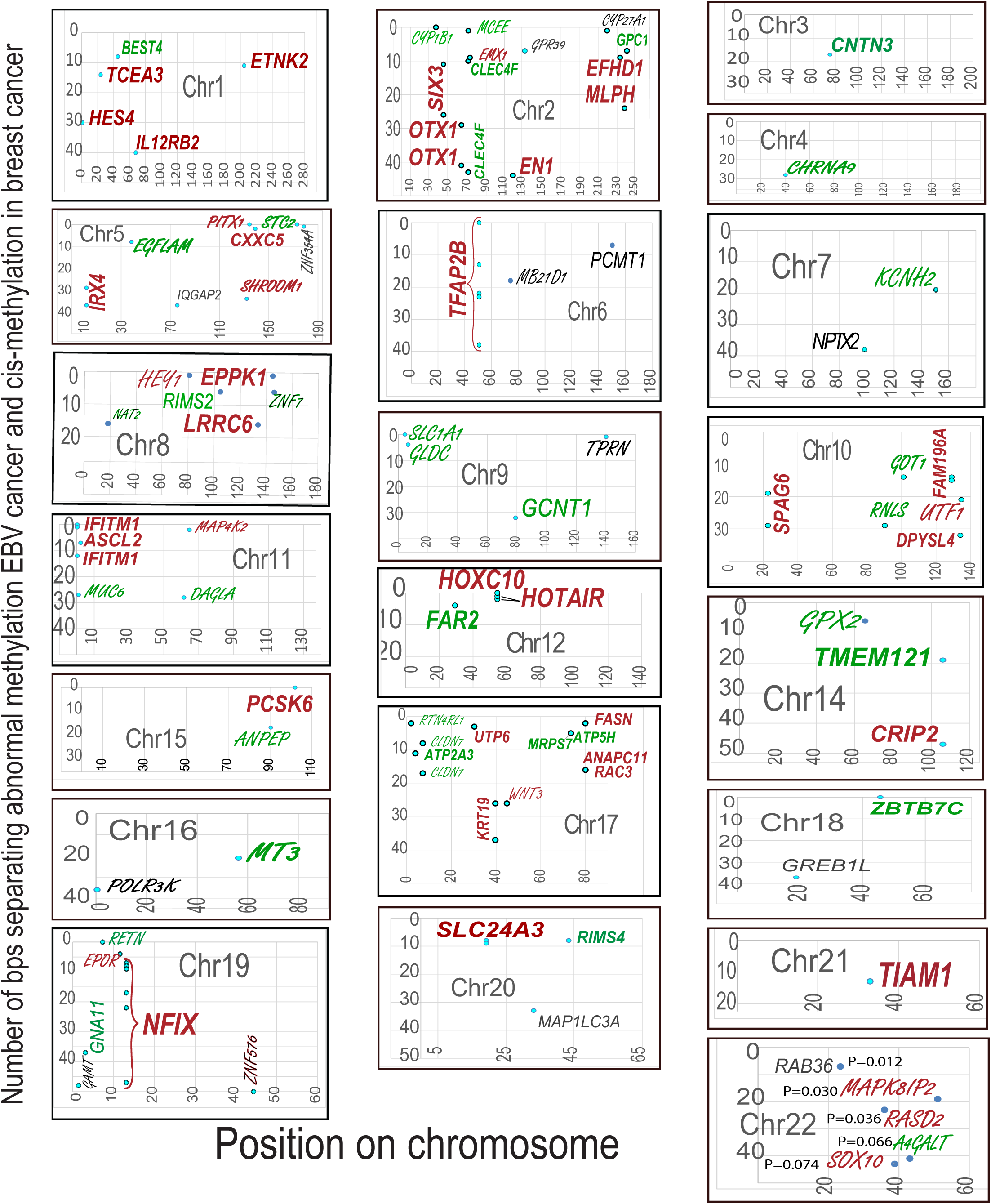
Some DNA gene control elements have abnormal cis-methylation in breast cancer that matches non-malignant oral keratinocytes that have recovered from an EBV infection. Positions that match either NPC or BL cancers were compared to abnormal methylation in breast cancers. The abnormal methylation sites focus on stem cell gene controls (red) and genes that affect the microenvironment (green). Other genes with matching methylated controls are indicated in black. These types of genes that are methylated in non-malignant oral keratinocytes but not in breast cancers are indicated by the script font. For chr22, p values show that the matching values include those that are unlikely due to pure chance.

Next, the exact genome coordinates were compared. As shown in Fig. 6 for cells from cured EBV infections, 14/22 autosomal chromosomes had methylation scars close to methylated loci that affected stem cell gene control in breast cancer. A few examples illustrate clear similarities. For chromosome 2, 20 of the 27 genes with abnormal methylation also had at least some relation to stem cells. On chromosome 12, the *HOXC* gene and *HOTAIR*, the *HOXD* controller are abnormally methylated, showing a clear relationship to the breast cancer stem cell data in Fig. 2. The *NFIX* gene on chromosome 19 had abnormal methylation in breast cancer, EBV cancer, and EBV infection. *NFIX* inhibition participates in both cancer and infection and may be essential for EBV pathogenesis. *NFIX* suppresses breast cancer cell proliferation by delaying mitosis through downregulation of *CDK1* expression. Eight chromosomes had new methylation loci that were still related to breast cancer stem cell gene controls (Supplementary Table S2).

These results show that EBV methylates non-malignant infected keratinocytes in much the same ways as it does in malignant breast cells even if the infection clears. Persistent methylation marks on stem cell gene regulators in non-malignant keratinocytes suggest that EBV associated methylation scars persist and are not easy to completely reverse.

### Control experiment shows abnormal methylation at sites essential to produce basal cell carcinoma of skin keratinocytes are not associated with EBV effects on non-malignant oral keratinocytes

Skin basal cell carcinoma (BCC) has no known relationship to EBV and was used as a control to test the association between methylation marks and EBV cancers in keratinocytes. Nearly 7000 hypermethylation sites in BCC [26] were compared to abnormal methylation sites in oral keratinocytes with a cured EBV infection. Fig. 7 shows that numbers of methylation sites in BCC (unrelated to EBV) near EBV-associated normal oral keratinocytes generally agreed less often than expected by chance. Using a 10 base pair window gave 1 match with 1.32 expected by chance alone. Assuming a Poisson distribution gives a p value of 0.73 (a 73% chance of seeing at least one match by chance alone). The p value rose to 0.27 at a 50 base pair window, which remained statistically non-significant.

**Fig 7.**
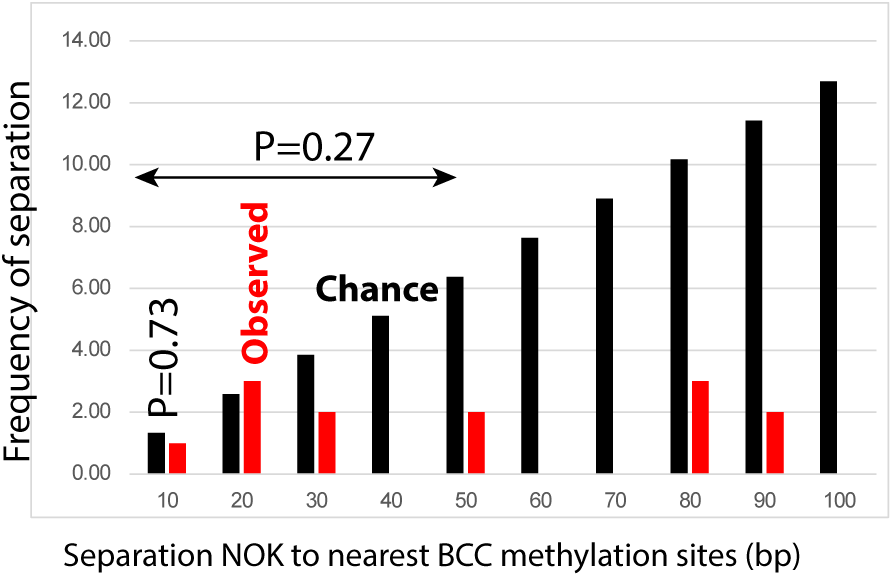
The frequency of methylated genes in normal oral keratinocytes that have recovered from an EBV infection (NOK)s within 50 bp of methylation sites in basal cell carcinoma of the skin is generally less than expected by chance.

### Breast cancer mechanisms of immune escape release EBV restraints to help explain why not everyone gets EBV cancers

EBV infects over 90% of people worldwide, but only a small fraction develop EBV-associated cancers. Variability in the activity of immune related genes may contribute to why only some people develop breast cancer. To test this idea, modification and expression of the genes related to immune responses were compared among the data for breast cancer patients.

As shown in Fig. 8A, 358 genes were near methylation sites in EBV cancers and 84 (23%) of the genes had functions related to the immune response. The 84 genes that impacted the immune system were mainly divided between those related to stem cells (40/84 genes) and those related to stem cell microenvironment (32/84 genes).

**Fig. 8A.**
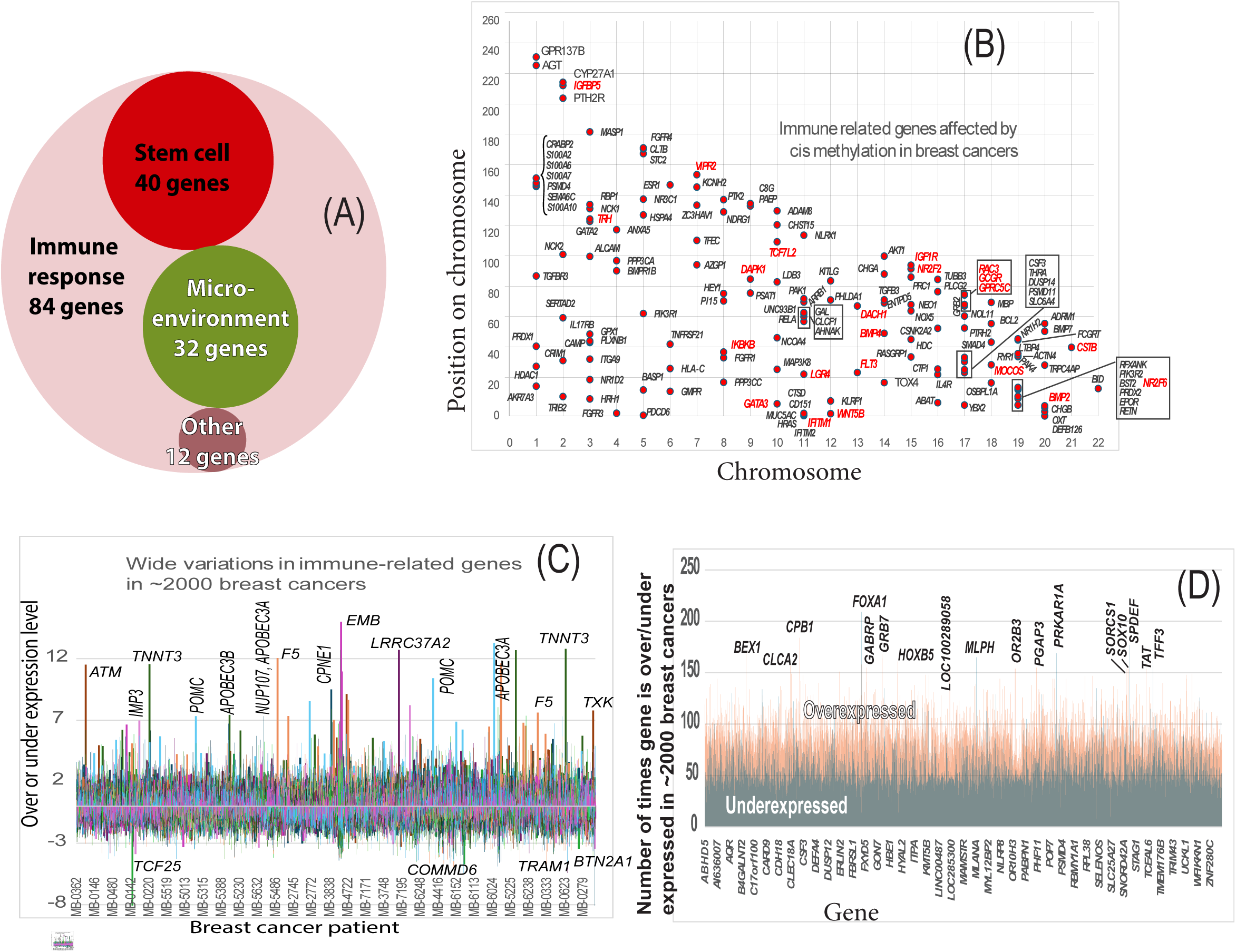
Control of genes in breast cancer by cis-methylation related to stem cells, the microenvironment, and other functions overlaps immune responses. This overlap occurs in about 23% of the cis methylated breast cancer gene controls near abnormal EBV methylation sites. Additional mechanisms of immune compromise are also characteristic of EBV infection. Fig. 8B. Immune response genes are affected by cis methylation in breast cancer. 4143 breast cancer genes under cis-methylation control include immune related genes on every chromosome that often cluster together. Fig 8C. Expression of immune-related genes varies widely in breast cancers APOBEC3B and 3A over-expression is common among breast cancers and both genes are significantly over-expressed in a few breast cancers. APOBEC3A and 3B are linked not only to breast cancer but also to EBV infection. Fig. 8D. Wide variation in gene expression in breast cancers is typical of all the breast cancers in the cohort. Only small percentages of the total 20603 genes are over or under expressed. This creates highly variable backgrounds for the development of breast cancers and contributes to explaining why not everyone with EBV infection gets breast cancer.

To further assess the influence of cis-methylation on immune-related genes, Batra’s 4143 autosomal genes controlled by cis-methylation [22] were compared to 1480 immunity genes [24] using METABRIC data from an ER+/PR-/HER2-subset of breast cancers. From the 4143 cis-methylation controlled breast cancer genes, 154 were related to the immune response in the subgroup and these are labeled in Fig 8B. Many of them are in tight clusters with 22 immune related genes near cis-methylation sites shared with EBV cancer chromosomes. If additional members of the same stem cell differentiation pathway are included based on a compendium of oncogenic pathways [46], the total rises to at least 50 immune genes. Of the original 22 immune genes, 19 were related to stem cell functions or conditioning the microenvironment.

To characterize potential consequences of these methylation anomalies, immune escape mechanisms were compared between ER+/PR−/HER2− subgroups in the METABRIC cohort and EBV-associated cancers [24]. Table 2 shows four obvious immune escape mechanisms in breast cancers that are also associated with EBV infections.

**Table 2.**
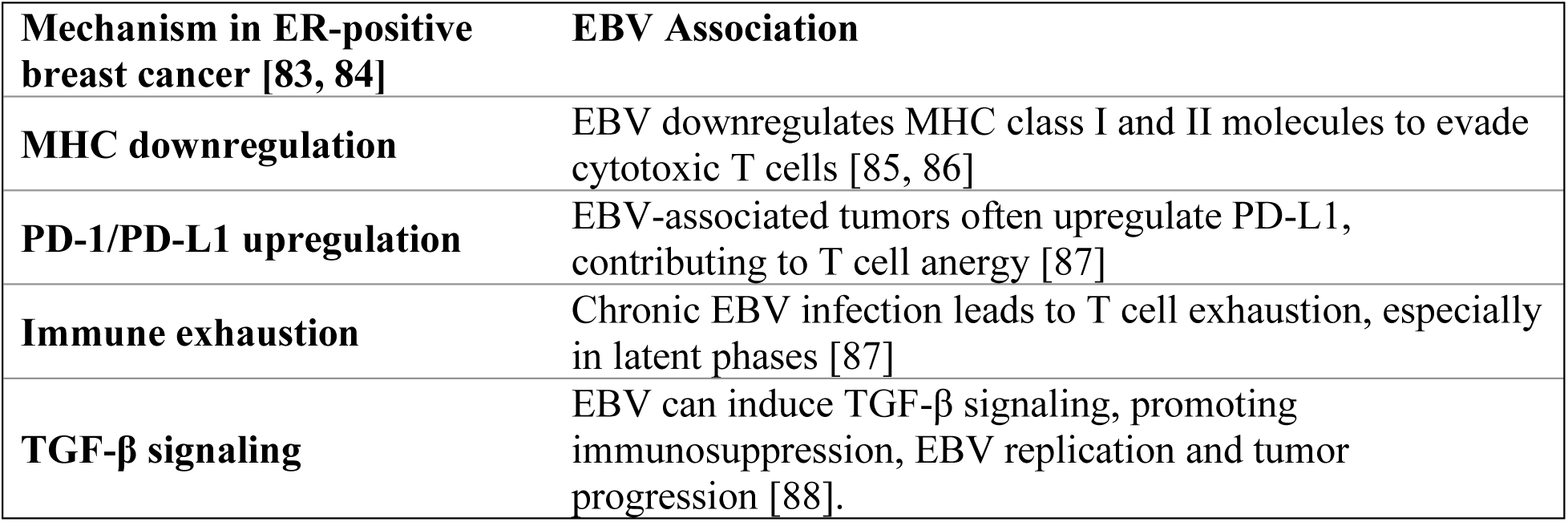
Immune escape mechanisms in ER positive breast cancers shared with EBV cancers.

In contrast 161 genes expressed in immune cells have been associated with breast cancer. These associations provide insight into tissue specific effects of gene expression[47]. The 161 immune related genes matched 97 METABRIC genes with expression values available and Fig 8B shows wide variation in their expression in different tumors. For example, as shown in Fig 8C, *APOBEC3A* and *APOBEC3B* are significantly overexpressed in a few breast cancers. The consequences of this overexpression have been linked to breast cancer tumor evolution, therapy resistance, and increased tumor mutational burden [48, 49]. Both *APOBEC3* genes are overexpressed in EBV infection and *APOBEC3A* is also overexpressed in malaria [50]. The two enzymes deaminate cytosine to uracil in single-stranded DNA, especially during DNA replication or repair. This leads to C-to-T and C-to-G mutations, especially in TCA and TCT motifs. Signatures correlating with ABOBEC overactivity are common mutations in breast cancer genomes [49].

These results show that methylation like that in EBV-related cancers is significant in shaping the immune response. The overexpression of *APOBEC3* genes is one force in a few breast cancers that extends the effects into oncogenic pathways. Immune imbalances are reiterated from EBV, further increasing chances that additional triggers will result in cancer. For example, chronic malaria exposure triggers BL in EBV-infected children. As shown in Fig. 8D, the genetic backgrounds for all these effects can vary widely. The approximately 20,603 genes measured in the cohort are over or under expressed in only low percentages of the ∼2000 breast cancers. These varying environments create different breast cancer backgrounds and contribute to explaining why not everyone with EBV infection gets breast cancers.

### The abundant EBV-like sequences in human chromosomes represent a small section of EBV DNA

To study the feasibility of a future vaccine against EBV, human chromosomes 3, 8, 11, and 17 DNA were arbitrarily selected for comparison to all known and available viral sequences. As shown in Fig. 9, many positions along the entire lengths of the four chromosomes resemble EBV and retroviruses, but the distributions of virus-like sequences differ. Chromosome 8 had the strongest resemblance to EBV with 6014 matching sequences (EBV homology scores over 500, indicating 355-500 identical bases). Because EBV-like sequences are so abundant on human chromosomes, EBV can mimic human sequences. The following five EBV-like human sequences have oncogenic properties in NPC [51]:

- *LMP2A* and *LMP2B* regulate the host aryl hydrocarbon pathway [52] and maintain latent infection.
- *A73* correlates with NPC occurrence [53]
- *BALF4* is a virion envelope glycoprotein in spikes that attaches to host cell surface proteoglycans. The viral and cellular membranes then fuse, enabling virus entry. *BALF4* dramatically enhances human cell infections [54]
- *BALF3* is an endonuclease that mediates mature virion production and packaging during the EBV lytic cycle[55]. The BALF3 protein causes double-strand breaks and micronuclei [56].
- *LMP-1* reprograms infected cells, preventing differentiation and inducing proliferation and inflammation [57]

**Fig 9A.**
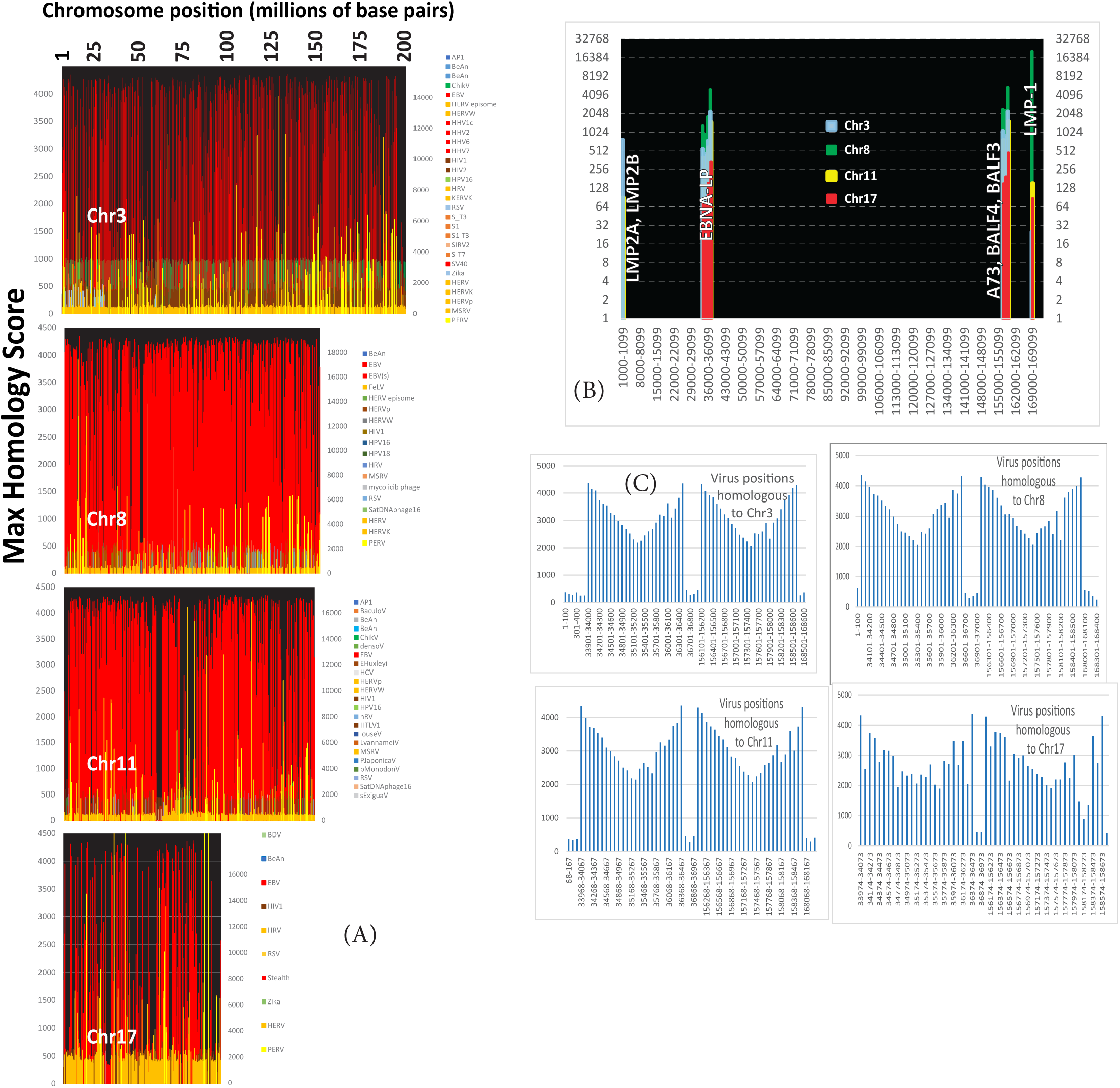
Positions on chromosomes 3, 8, 11, and 17 with significant DNA sequence homology to any known virus. Fig 9B. Genes on the EBV chromosome that correspond to the positions of homology in Fig. 9A. Fig. 9C. Number of homologous sequences at each position in the EBV genome.

This result shows that thousands of full or partial copies of EBV-like sequences relate to human genome instability (Figs. 9A, B, C) and emphasizes how dangerous EBV infection can become. Fortunately, human DNA only matches about 2500 bases within the 173,000 bases in EBV genomes. Only a few of all possible EBV genetic regions probably exist in the human genome suggesting that the products of these regions should be excluded from vaccines or treatment protocols.

## Discussion

Methylation of breast cancers focuses on the same stem cell gene control elements that are targeted in EBV-cancers and even in cleared EBV infections. EBV changes the accessibility and expression of genes encoding critical transcription factors activated across the entire human genome. For instance, abnormal methylation shared with EBV cancers was common for *HOX* genes and their interactions with morphogen and morphogen regulated pathways. These HOX interacting routes included Wnt-beta catenin, *BMP, TGF, FGF*, Retinoic acid, *Hedgehog*, *NOTCH* and *Hippo.* Abnormal methylation of *HOX* genes can promote cancer not only by confusing tissue organization, but also by increasing their oncogenic activity and interfering with their roles as tumor suppressors. At least one *HOX* gene has been correlated with breast cancer outcome [58]. Genome safeguards disabled by EBV are characteristic evidence of prior EBV infection, regardless of whether the infection is still active [5]. The epigenetic changes that match EBV infected cells in the current work might account for some of the similarities in chromatin breaks found between breast cancers and EBV cancers [5].

The current results probably explain why stem cells or their lineage descendants are absent in normal adult human breast basal cells but apparent in breast cancers. The virus causes persistent changes in stem cell gene control elements and this produces or selects for cancer stem cells. These stem cells are friendly to viral replication and labile to malignancy.

Direct model experiments indicating that EBV infection affects breast stem cells have already been published [59]. The initial experiments engineered mammary epithelial cell lines (MECs) with hTERT, SV40 large T-antigen, and Ras V12 oncogene. EBV marked with the gene for green fluorescent protein cooperated with the Ras oncogene to multiply early MEC progenitors that had a stem cell phenotype and a differentiation block. The ability to form mammospheres increased up to ten-fold and was used to count stem cells. The infected cell lines were then introduced into immunodeficient (NOD / SCID) mice. The emergence of triple negative mammary tumors greatly accelerated in the immunodeficient mice if the cd21 receptor was present [59]. Although there are potential objections to the methodology, organoid culture assays, genetic lineage tracing in human cancers, and in vivo microscopy of mouse breast tumors have all supported the initial xenotransplantation experiments [60].

Additional published laboratory results also support the current findings. Single cell transcriptomic profiling of breast cancer found a differentiation depending on signals from the microenvironment and diversity in mesenchymal differentiation states, including stem cell markers [14]. The EBV protein LMP2A is capable of inducing stem like cell populations in NPC [61]. EBV is also known to infect hematopoietic stem cells giving rise to a wide variety of different infected lymphoid and myeloid cell types [62]. EBV protein LMP-1 binds the *CEBPA* enhancer and induces dedifferentiation [10]. *CEBPB* is the *CEBPA* binding partner in a heterodimer. *CEBPB* control regions on chromosome 20 were differentially methylated in breast cancer and corresponded to EBV cancers. *CEBPB* is essential for differentiation in hematopoietic stem cells, muscle and skeletal stem cells.

EBV reprogramming of its host’s epigenetic system has been observed in NPC where there are characteristic and massive changes in methylation [16, 17]. NPC has hypermethylation in at least 80% of cases [18]. In the remaining 20%, subtle changes in specificity and activity of methyltransferases DNMT3A and DNMT3B may account for methylation deficits [63]. EBV suppresses the removal of methyl groups by demethylation pathways, which also alters chromatin exposure [18]. Like NPC, BL causes epigenetic reprogramming by silencing at least one critical transcription factor [64]. Both NPC and BL have forms in which EBV is undetectable, compatible with loss of virus as the cancer progresses.

Epidemiologic studies reported EBV infection was about five-fold more likely in breast cancer tissues than in non-malignant controls [65], but the virus can disappear [66]. However, EBV still leaves evidence of infection behind even in non-malignant cells. EBV-infected oral keratinocytes that expressed telomerase have hypermethylated CpG islands as scars that normal cells do not have. The methyl groups remain even if the virus is lost during repeated host cell divisions. These epigenetic alterations change gene expression, interfere with differentiation, and can lead to an invasive phenotype [45, 67]. Breast cancer methylation was observed near the same positions. This persistent methylation interferes with differentiation in keratinocytes and does not occur in never-infected cells [45]. More broadly, the implication is that a virus infection can change the human genome, even if the virus does not integrate its DNA. Many of these changes are deleterious and support wider use of vaccination against virus disease. Basal cell carcinoma which is unrelated to EBV is also unrelated to persistent methylation in keratinocytes.

Gene therapy or genetic manipulation of stem cells is a promising general strategy for treating many human disorders. The results here show that stem cells are susceptible to damage from infection and require at least an epigenetic quality assessment before use.

The significant homology between the human and EBV genomes complicates the development of a vaccine or treatment protocol. The results presented here and previously [5] predict that vaccination against near-universal EBV infection could significantly lower the incidence of breast cancer.

## Methods

### Breast cancer patient data

All breast cancer data came from female patients with primary breast cancers. Original publications [35] and Kaggle are freely available sources used to obtain clinical and genome data from the METABRIC cohort of 1904 patients. Briefly, 1459 patients in the cohort had tumors classified as ER positive breast tumors, and 445 as ER negative. Grades 1, 2 and 3 were in 165, 740, and 927 patients respectively. Patient ages ranged from about 22 to 96, with most (1200/1904) between ages 47 and 72. The cancers were mostly ductal (1454 patients). The original publications also described tissue collection procedures, clinical data, gene expression profiles, copy number aberrations, and point mutations [35, 68]. The publications also provided stage, grade, tumor size, and the status of lymph nodes, ER data from in situ hybridization, PAM50 gene signature [69], and breast cancer driver genes [22].

### Breast cancer methylation data

Using samples from the METABRIC cohort, published breast cancer differential DNA methylation positions had been derived [22] from whole genome reduced representation bisulfite DNA sequencing of 1538 breast tumors and 244 adjacent normal samples. Identification of methylation sites in breast cancers that correlated with gene expression through promoters and enhancers on the same chromosome was taken from published results [22]. Briefly the published identification procedure screened breast cancer genomes for Pearson correlation of promoters on the same chromosome as methylation sites. Large methylation (“trans”) effects that could effect gene controls on multiple chromosomes were eliminated. If a known gene promoter had the strongest, most negative correlation with its corresponding gene, it was taken as a specific (“in cis”) effect. Correlations were all based on >50 samples with FDR’s limited to <0.05). Non-promoter cis-regulation candidate searches did not depend on a one to one correspondence between methylation sites and regulatory targets. Since a locus can correlate with multiple genes on the same chromosome, correlation values for every locus within 500 kb of any of the genes were ranked. The genomic distance (in bp) was then calculated between each methylated locus and the coordinates assigned to its expression profile. A locus was defined as paired with a gene control region if the transcription start site of its most strongly correlated gene expression was within 500 kb. Shuffled data was used to estimate the probability that the pairing occurred by chance. The ratio between pairing events in real vs shuffled data was then taken as the false discovery rate [22]. This protocol identified 4143 genes on the 22 human autosomal chromosomes as regulated by sequences on the same chromosome as the regulated gene [22].

### NPC patients and methylation data

NPC comparisons to breast cancer were based on unrelated and independently published data [18] for NPC biopsies from 15 sporadic NPC patients and 9 non-tumor adjacent tissues. Their cancers were all primary NPC’s at stages II (3 patients), III (7 [patients) and IV (5 patients), with 11 patients surviving. Ages ranged from 38 to 82, composed of 13 males and two females. Differential methylation in NPC was obtained from published whole genome bisulfite sequencing data reported as the average methylation difference between NPC and normal tissue. The results showed 16,910 differentially methylated stretches of DNA had average methylation that differed from the same region in normal DNA by an absolute value of at least 0.2. From these 16,910 regions, 6036 were <500 bp long with average differential methylation of –0.336 to 0.463.

Three patient samples were globally hypomethylated and 12 were globally hypermethylated. An independent patient cohort of 48 NPC patients had been used to validate the whole genome bisulfite sequence results [18]. For analysis technical duplicates minimum difference calculations and were not considered.

### BL patients and methylation data

BL comparisons to breast cancer came from published data for 13 BL patients [19], ranging in age from 3 to 18. Eleven patients were male and two were female. Tumors in all the patients had breaks in the *MYC* gene and *IG-MYC* translocations. All the tumors had differential hypermethylation that correlated with gene expression. Over two-thirds of the differentially methylated regions were <500 base pairs in length but the lengths ranged from 8-5160. Reference DNA came from the non-neoplastic germinal center B-cells of non-neoplastic tonsils removed from four patients, ages 13-30. The data used in the current work began from 42,381 differentially hypermethylated regions on the 22 autosomal chromosomes. The average methylation in germinal center B-cell DNA segments was subtracted from the average rate in BL and the values were limited to a difference of at least 20% across DNA region that were differentially methylated. The genomic positions of all these regions were compared to genomic coordinates of cis differential hypermethylation sites that affected breast cancer gene controls.

### EBV infected normal cell data and transiently infected normal cells

The published data [45] used came from a clonal population originating from normal oral keratinocytes (NOK) that had been immortalized by human telomerase (hTERT) [45]. Briefly, the original publication had infected NOKs by coculture for 24 h with anti-IgG-induced Akata BL cells. After removing B-cells, infected cells were selected for antibiotic resistance on 10 passages. Clones with transient infection were produced by removing antibiotic selection pressure for ten additional passages, after which flow cytometric single-cell cloning was performed. EBV encoded RNA (*EBER*) in situ hybridization confirmed the EBV positivity of infected and transiently infected cells. Uninfected parental cells and plasmid-transfected controls were cultured under identical conditions; infected cells were subjected to single-cell sorting. DNA fingerprinting analysis authenticated the identity of cell clones in relation to uninfected parental controls. Methylation was determined by reduced representation bisulfite sequencing for six samples: uninfected parental cells, control cells transfected with the plasmid vector used, one EBV-positive clone, and three clones transiently infected but EBV-negative [45]. Although the age of the donor was not specified, normal oral keratinocytes typically come from adult donors. For example, a well-known hTERT-immortalized oral keratinocyte line is OKF6/TERT-1, derived from the oral cavity of a male donor, age 57 [70].

### Skin basal cell carcinoma methylation data

Basal cell carcinoma data (BCC) came from 16 BCC biopsy specimens from 11 women and 5 men, ages 55-89. The samples were 8 nodular BCCs, and 8 sclerodermiform BCCs [26]. Tumor sites included face, shoulder, head, ear, lip, nose, and pectoral areas. Data had been mapped against the human reference genome (i.e., hg19). Whole genome methylation profiling was with Infinium MethylationEPIC BeadChips.

### Calculations of matching abnormally methylated positions

Distance between abnormally methylated positions in breast and EBV cancers were calculated as the minimum value obtained by subtracting start, end, and mid-points of differential methylation in NPC [18] or BL [71] from these values for cis hypermethylation [22] in breast cancer. The same piece of DNA could be abnormally methylated at different positions. A strict cutoff of 20% difference from normal cells was used over the entire length of DNA segments being compared. An example of a typical formula used to calculate minimum distances is as follows. =MIN(ABS(D2-$K$2:$M$1532),ABS(E2-$K$2:$M$1532),ABS(F2-$K$2:$M$1532)), where values in column D through F contain breast cancer cis hypermethylation positions (start, end, mid-point) and values in columns K through M contain differential hypermethylation positions (start, end, mid-point) in EBV cancer (either NPC or BL). The Poisson distribution was used as the standard model for estimating the probability of a given number of rare, independent events occurring in a fixed space or time. The p-value was calculated from Poisson distributions as follows: 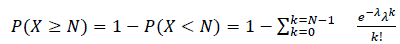. Where λ = the number of matches expected from random events and N=the maximum number of matches found. Poisson distributions were evaluated using a Python 3.13 script in a Jupyter notebook within the Anaconda3 platform.

For calculating the control breast cancer vs basal cell skin carcinoma beta values of >=0.5 were used. Beta is defined as methylation sites divided by the total of methylation and non methylation sites.

To test for problems caused by including excessively long DNA sequences, values of differential methylation were cut off in controls at 1000 bps, making only minor differences in the results. DNA sequence positions all came from reads that had all been positioned according to Human Genome Assembly GRCH37 (UCSC Release hg19). These alignments were used directly from the original publications and not corrected or manipulated in any way.

### Calculation of frequency of close agreement

Frequencies of agreement between breast cancer and EBV cancer were calculated after normalizing the numbers of NPC differential methylation sites to the same total numbers of cancers as the BL data (15 NPC vs. 13 BL). Frequency calculations were performed in Excel.

### Relationships to stem cells and stem cell microenvironments

Stem cell associated genes were defined as those essential for self-renewal, asymmetric cell division, terminal differentiation from a pluripotent state, or regulators of these processes. In traditional usage, stem cell genes refer to transcription factors and other regulatory molecules—such as *OCT4, NANOG,* or *SOX2*—that maintain pluripotency and self-renewal. In the current work, morphogens and other transcription factors that define cell fate and lineage were also included. Mere expression in stem cells was not considered sufficient for this classification.

The functions of about 350 genes controlled by abnormal methylation at <50 bps separation distances between breast and EBV cancer or EBV infected non-malignant cells were determined by manual curation. Artificial intelligence programs such as Microsoft Copilot and Google provided valuable assistance; however, both occasionally produced errors. Accordingly, all outputs were carefully reviewed, verified, and validated with literature references. Each breast cancer abnormally methylated gene control site within <50 bp of an EBV related methylation site was initially queried for a relationship to stem cells, to regulating stem cell gene expression, differentiation fate, or use as a stem cell marker. Stem cell databases were included and routinely reviewed [25–28]. The primary criterion for identifying a gene as a stem cell related gene was a credible reference documenting this classification. At this stage of the work, embryonic, pluripotent, and multipotent stem cells were not considered separately.

Accompanying searches for some relationship to the stem cell microenvironment were also conducted for matching breast cancer-EBV cancer methylation loci. Genes that influence the microenvironment were taken as those that code for proteins involved in extracellular matrix remodeling, transport factors, structure surrounding the cell, or other modulatory factors. Changes in internal cellular conditions also can directly determine how cells sense and respond to their microenvironment. In contrast some genes were characterized as “Other” (not stem cell genes) because their roles in cell structure or metabolism only indirectly affected stem cell gene products or the stem cell microenvironment.

### Breast cancer gene expression data

Breast cancer gene expression data came from the METABRIC study [35]. The study design had two cohorts: Discovery (997 patient samples) and Validation (995 patient samples). An Illumina HT12 v3 microarray was used to determine gene expression levels. Expression values were presented as Log2(transformed intensity values). To permit cross-sample comparisons, Log2 values had been normalized to z-scores. Z-scores for gene expression that were at least two standard deviations from the mean give a p value of <0.05. Expression of genes that had these z-scores were then compared to stem cell genes with abnormal methylation positions that were <50 bp from methylation positions in EBV cancers.

### Calculation of overlap in immune related methylation positions in breast cancers vs methylation position in EBV cancers

An Excel formula was created to check whether the value in cell **K2** (representing an EBV cancer methylation position) falls within a reported range from start to end for any of three sets of methylation positions in CGI regions for breast cancer immune prognosis genes (in rows 2, 3, and 4 across columns AB to AF).

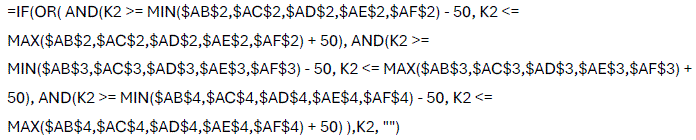

### Comparisons of distributions calculations

Tests for departure from normal distributions and other statistical calculations were made using the StatsDirect statistical program. These tests routinely showed that hypermethylation and breakpoint positions on any chromosomes were unlikely to be normally distributed. Therefore, Mann-Whitney nonparametric tests were used to compare the distributions of differentially methylated positions. Correlation between epigenetic modifications on breast vs EBV cancers was by linear regression analyses with a power for 5% significance of 99.99%. Kolmogorov-Smirnov and Spearman’s tests were also used. Frequencies were plotted to verify the distributions of coinciding areas.

### Window sizes to compare methylation sites in breast and EBV cancers

Window sizes of <50 were used to compare differential methylation sites in breast cancer and EBV cancers. This value is consistent with a minimum typical length for enhancer DNA (50-1500 bp). The 50-base comparison at start, end, and middle regulatory sites fully covers <200 bps and thus finds only the minimum number of identities from longer differentially hypermethylated regions. The stringency of the <50 base window size was checked by determining how methylation sites were distributed with respect to transcription start sites. The frequencies of methylated sites at –500,000 to +500,000 distance from the published [22] paired gene transcription start sites were calculated from the pooled data for human chromosomes 1-22. Calculation of the frequencies of separation between differential hypermethylation and transcription start sites for breast cancer chromosomes was done to assess the rigor of a window size of 0 to <50 bps. This test revealed 927/4143 methylation sites within 1000 bases, 261 were within 500 bases, one within 200 bases, and none within fifty bases. Based on this result, the 50 base pair comparison window was judged to be rigorous and was therefore used.

### DNA sequence homology

Homology between human and viral DNA was determined using the NCBI BLASTn program (MegaBLAST). E values less than 1e-10 was the criterion indicating significant homology.

## Conclusions

These findings illustrate a selective pressure from a viral infection that acts to facilitate malignancy in cells with stem like properties.

1. **EBV’s Role in Breast Cancer**: Epstein-Barr Virus (EBV) infection produces or selects for stem cells that permit viral replication and allow malignancy. Abnormal methylation sites in breast cancer closely match those in EBV-associated cancers, suggesting a common mechanism driven by EBV. These findings explains why stem cells are absent in normal breast basal layer but present in breast cancer.
2. **Impact on Stem Cells**: EBV targets and disrupts the regulation of stem cell-related genes. Reshaping the host cell genome and epigenome creates a cell friendly to viral replication and susceptible to malignancy.
3. **Persistent Epigenetic Changes**: Even after EBV infection clears, the virus leaves lasting methylation scars on non-malignant cells. These scars affect some stem cell control sites, occur in cancers, and may push cells toward malignancy.
4. **Vaccine Potential**: The significant homology between human and EBV genomes complicates vaccine development, but the study identifies specific EBV sequences to exclude from immunogens. A childhood vaccine against EBV could help preserve normal stem cell functions, potentially reducing the risk of breast cancer.
5. **Therapeutic Implications**: Understanding EBV’s impact on stem cell genes is crucial for protecting stem cells and for quality control of regenerative medicine.

## Data Availability

Datasets used are well-documented and freely available from the original sources or the corresponding author on reasonable request.

---

**Supplementary Table S1.**
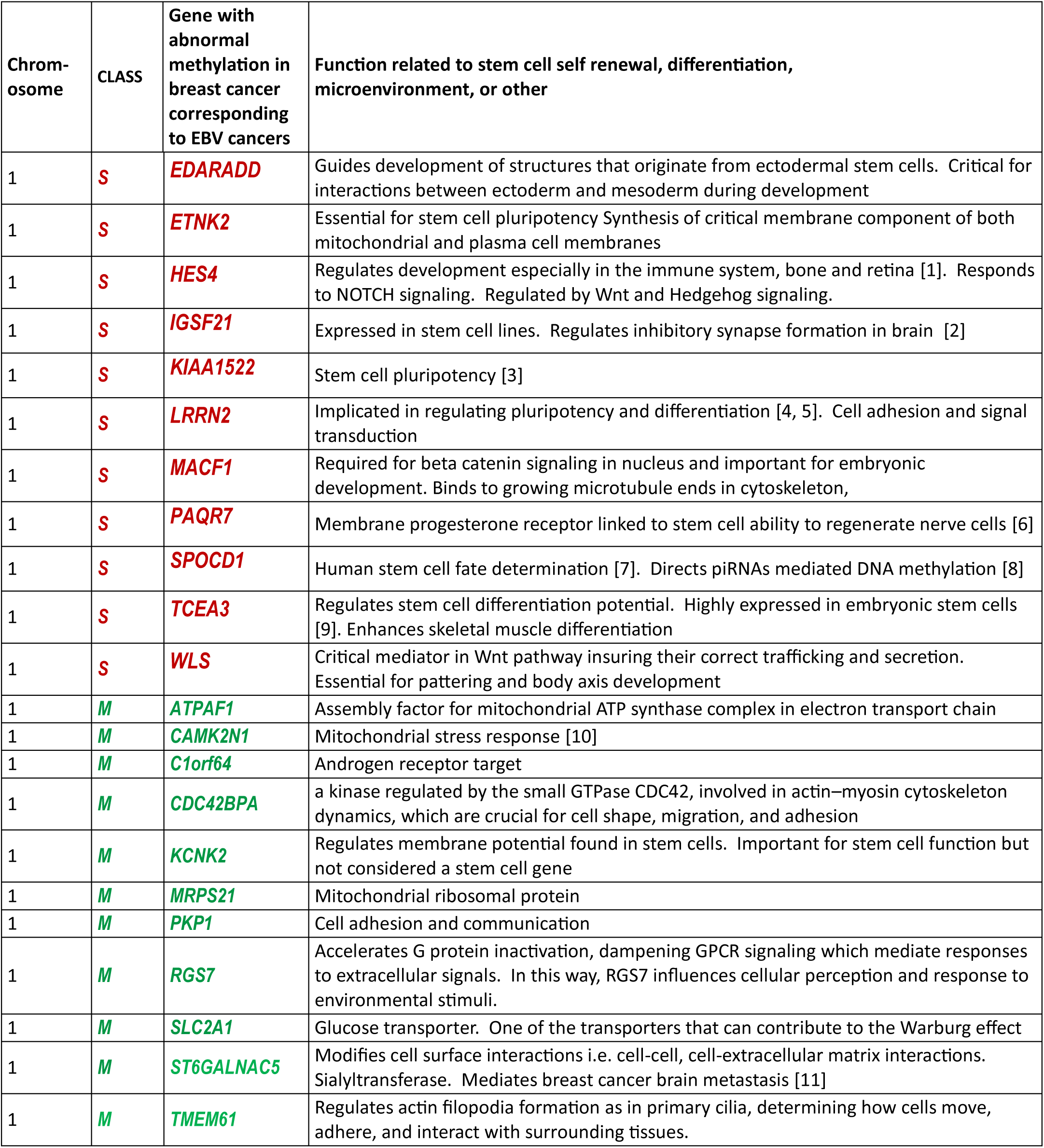
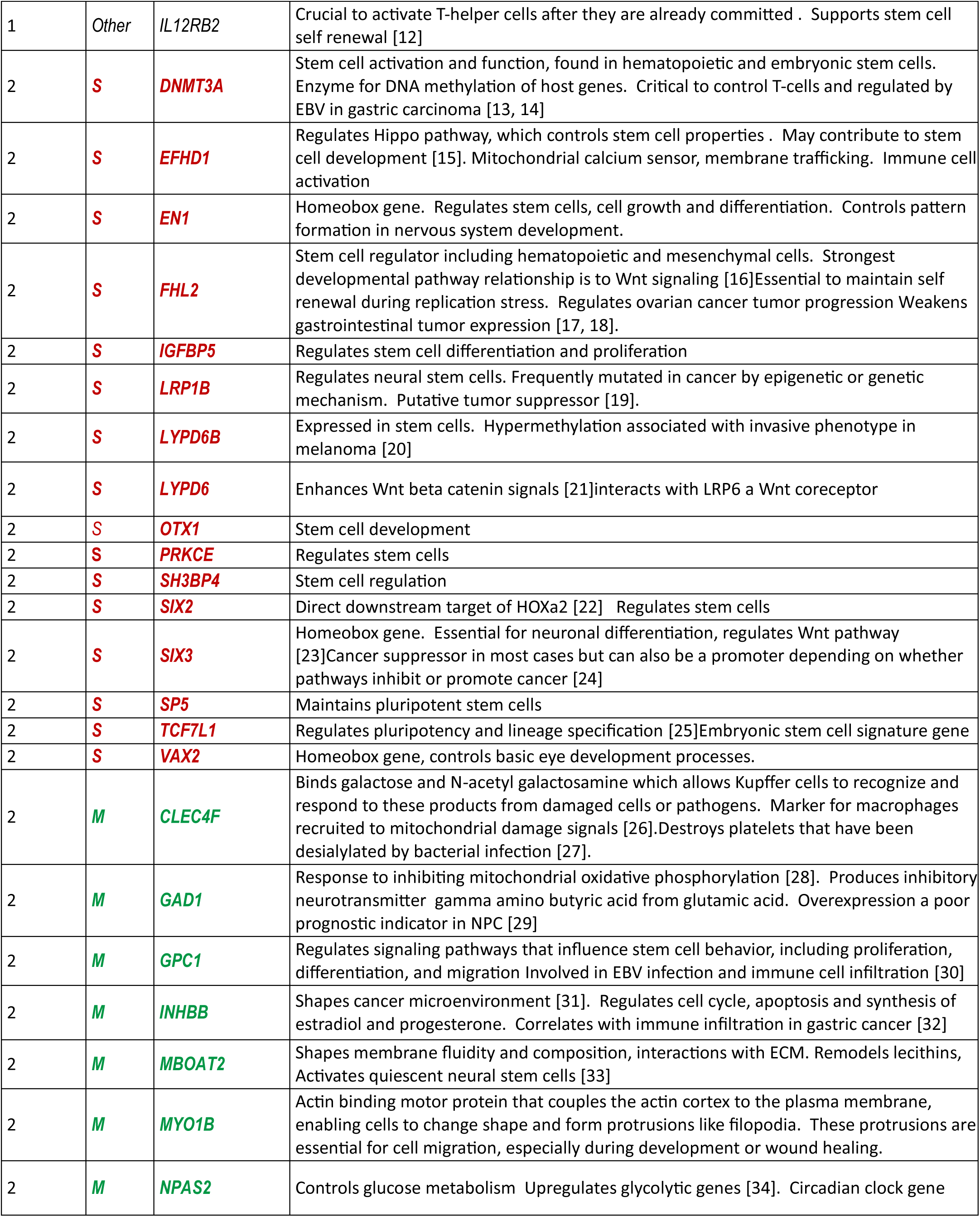
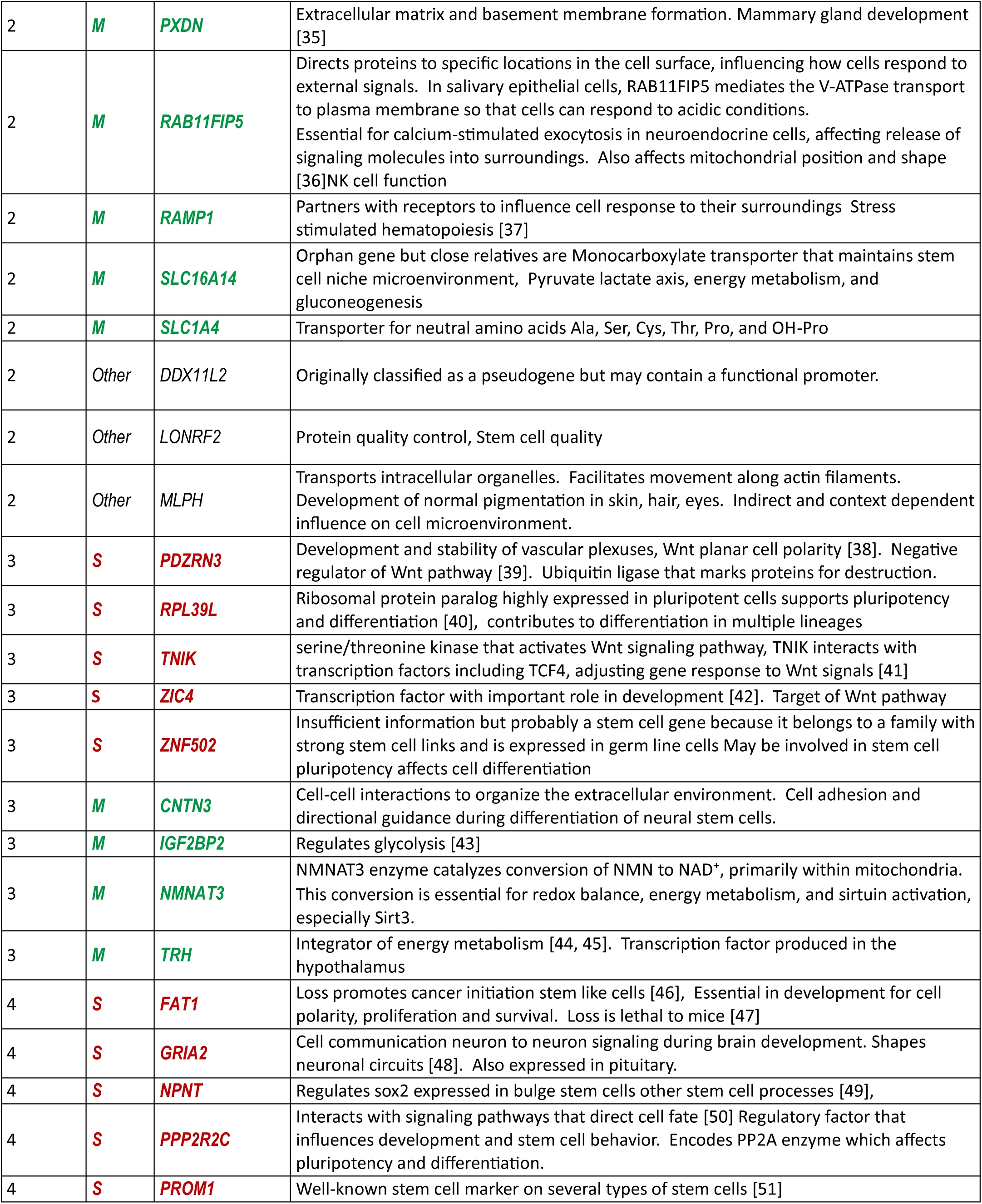
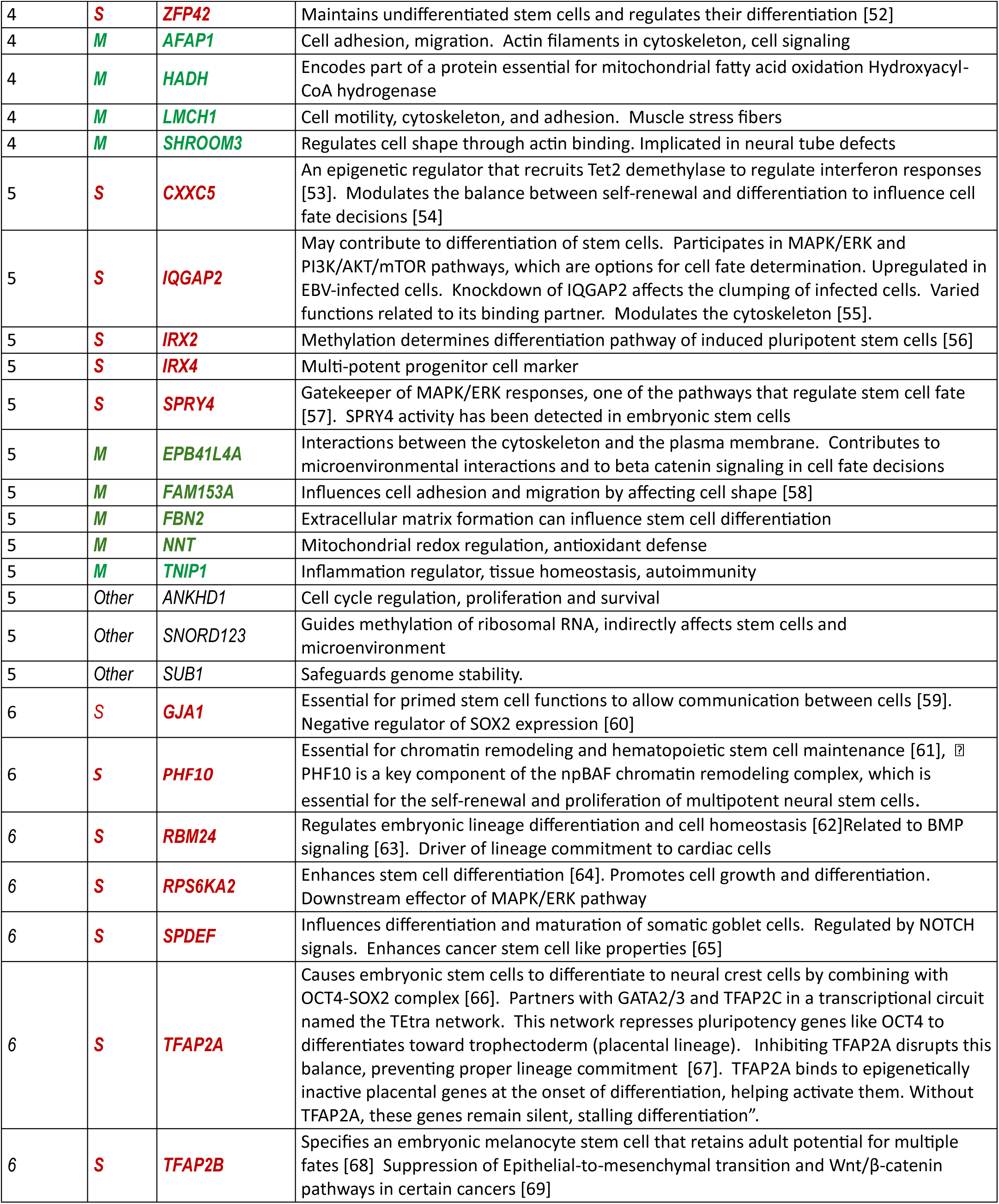
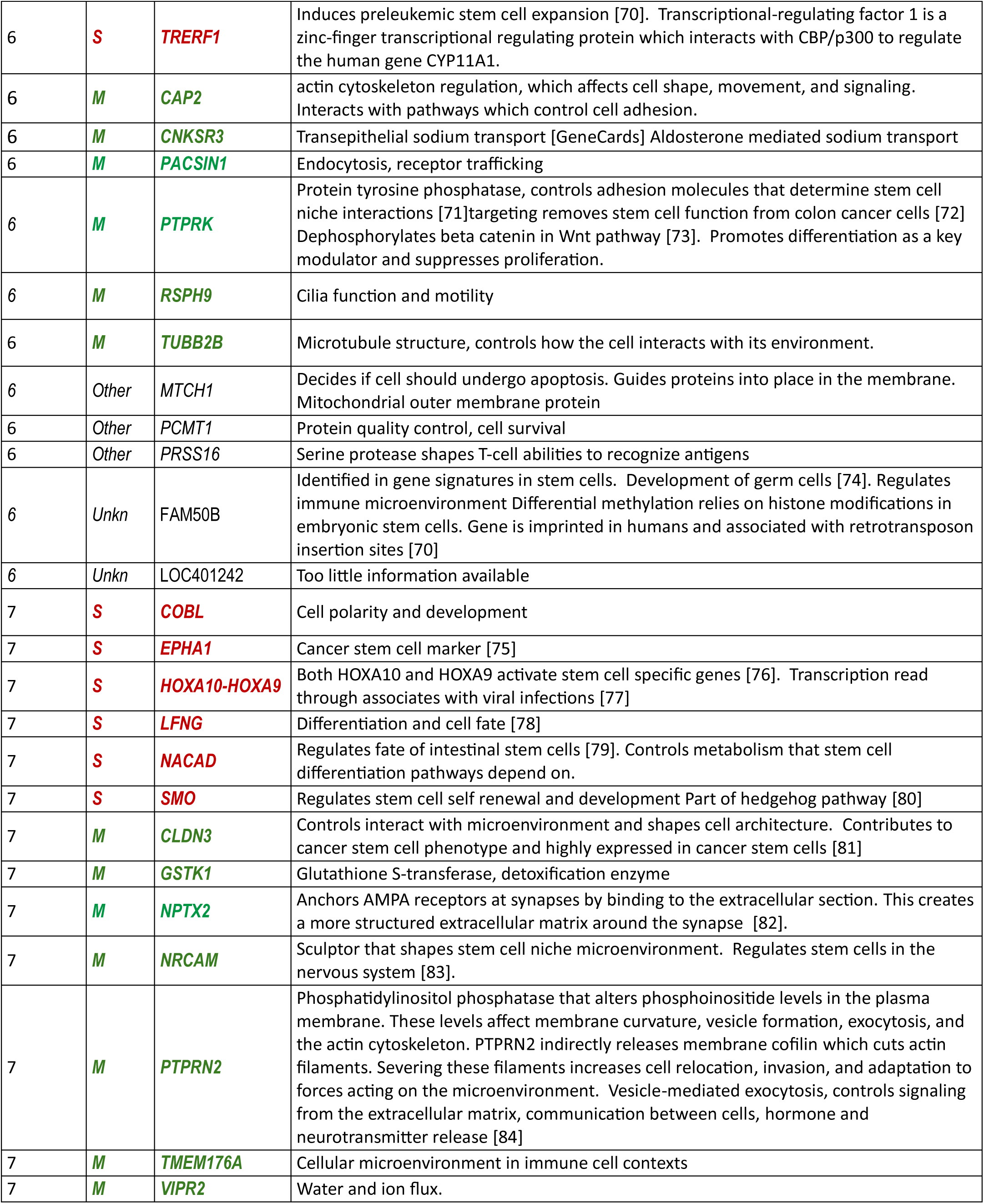
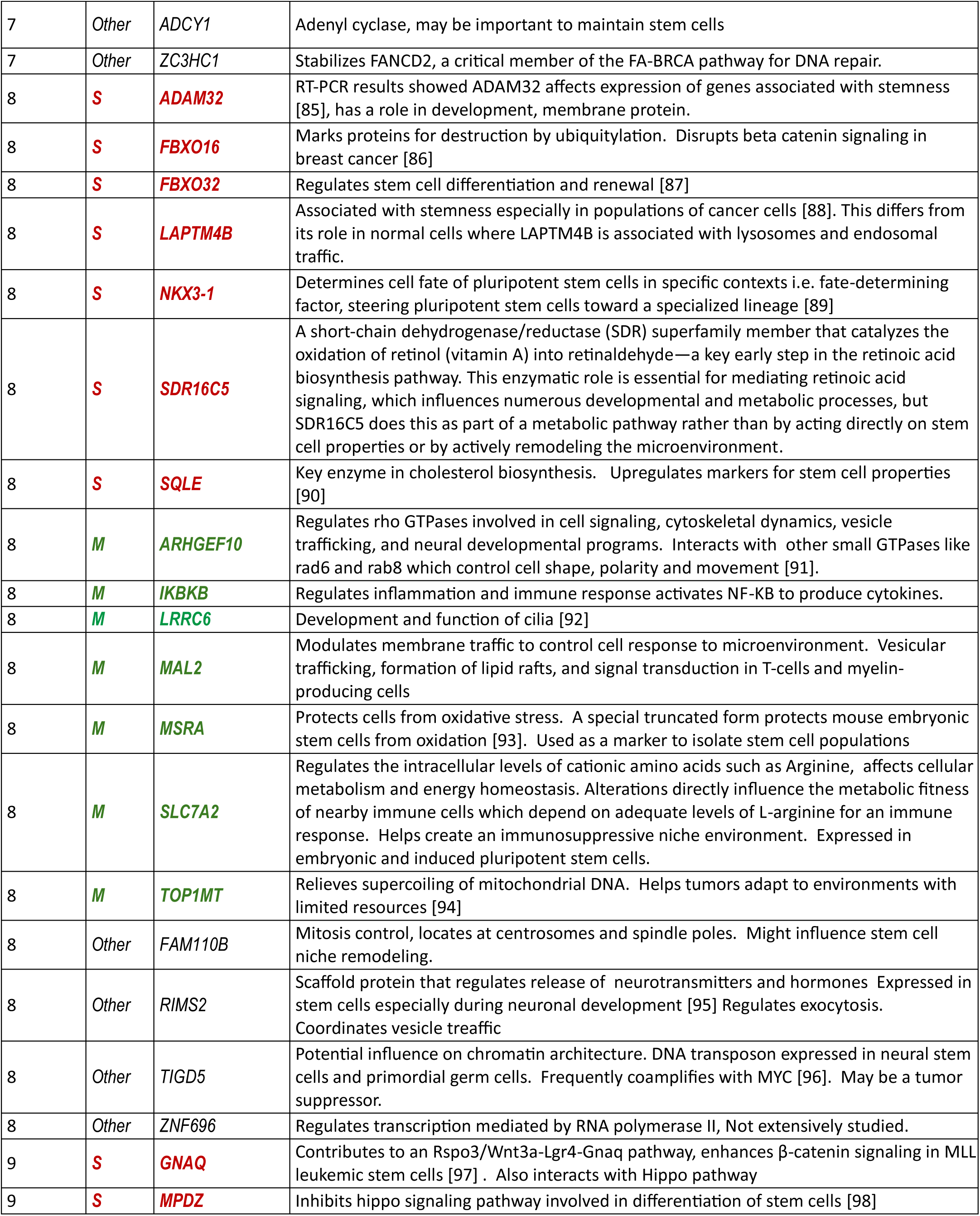
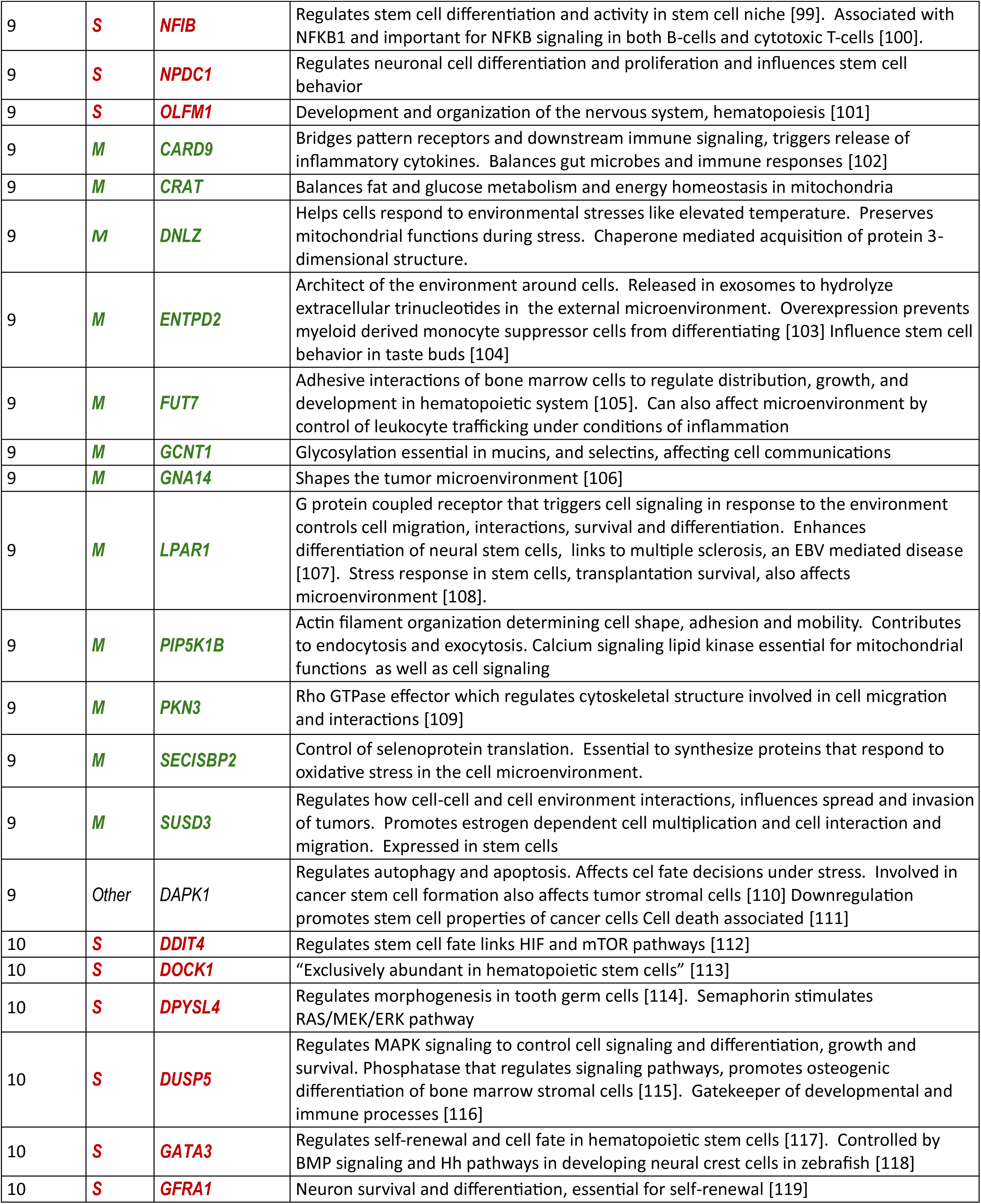
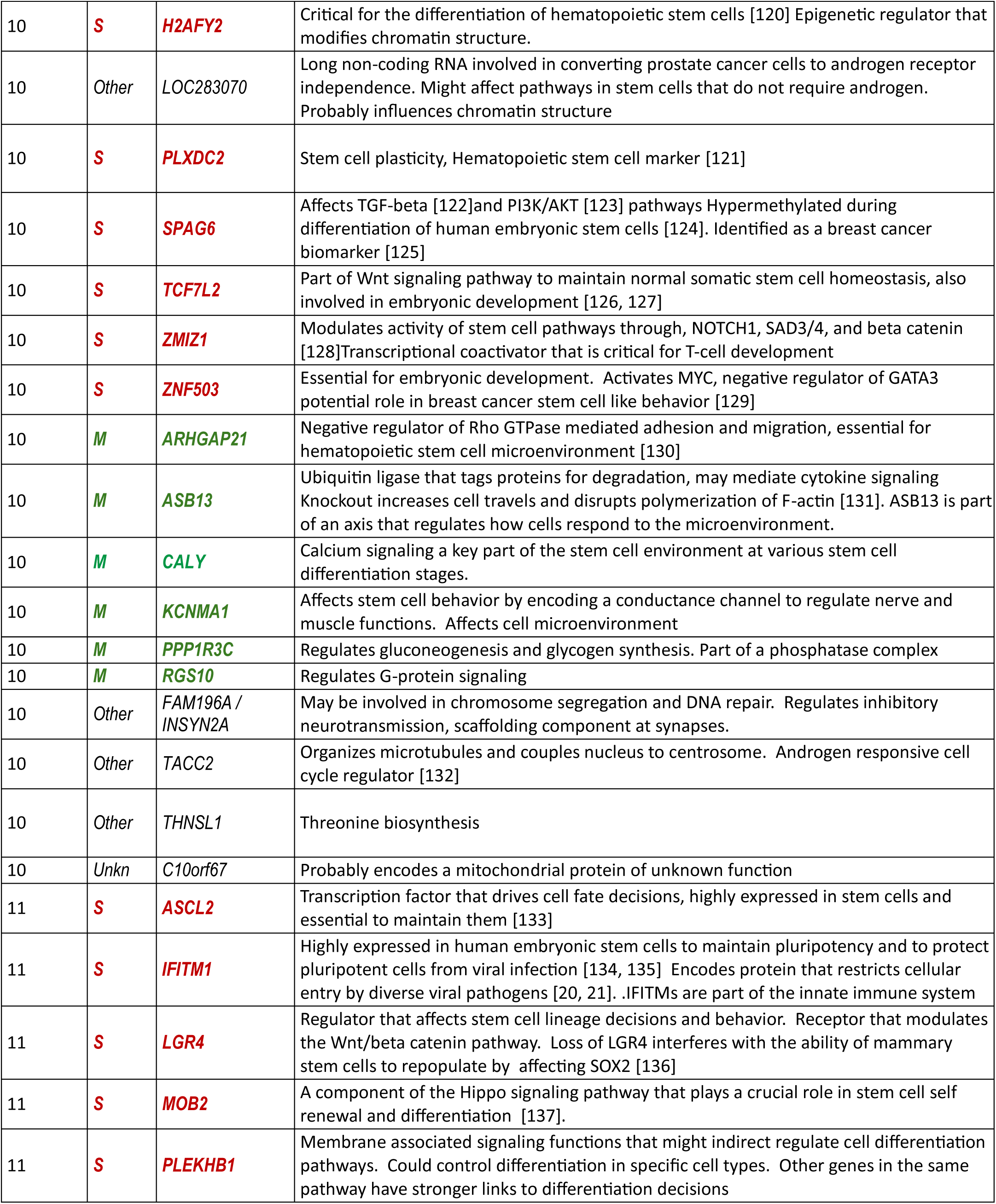
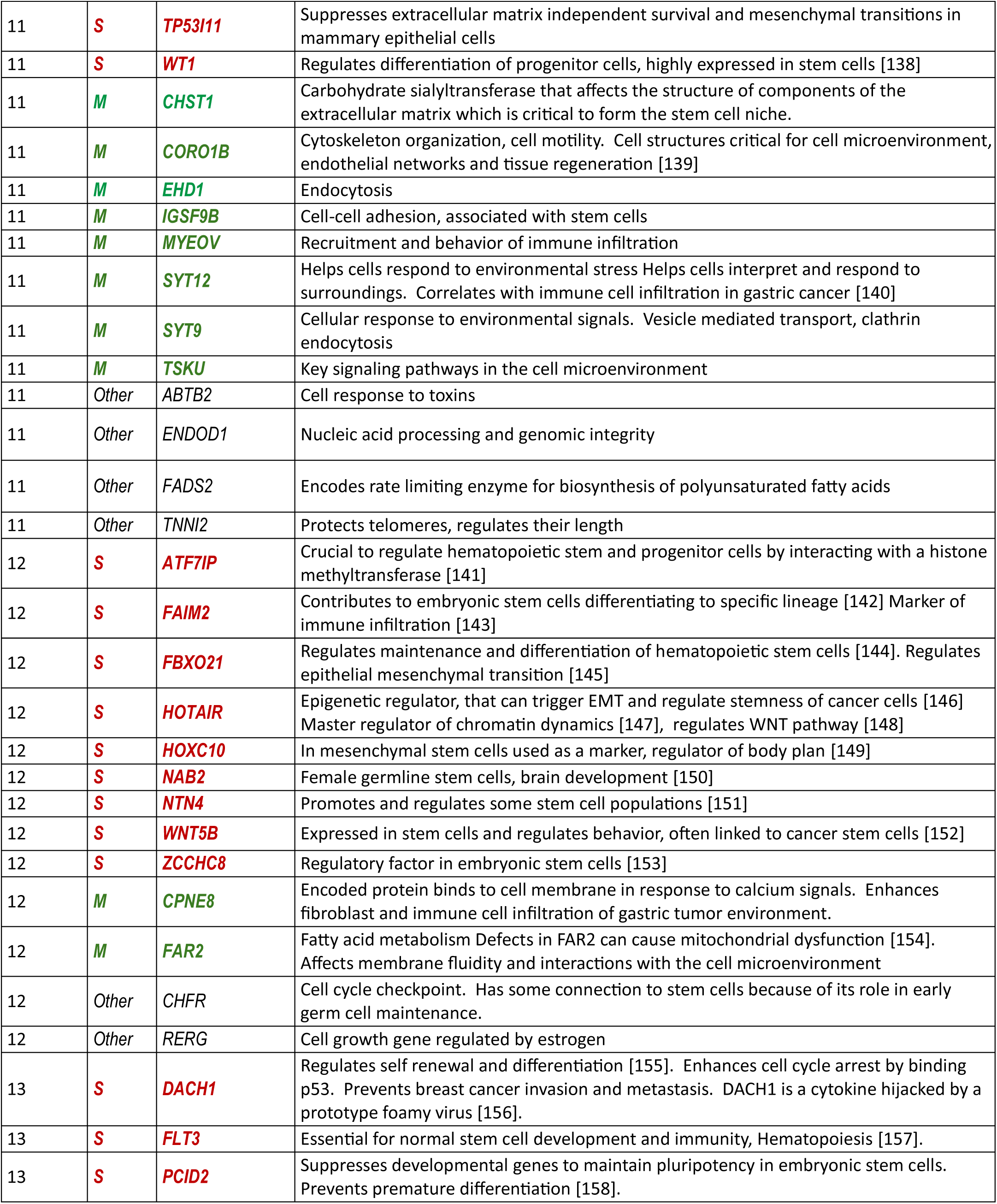
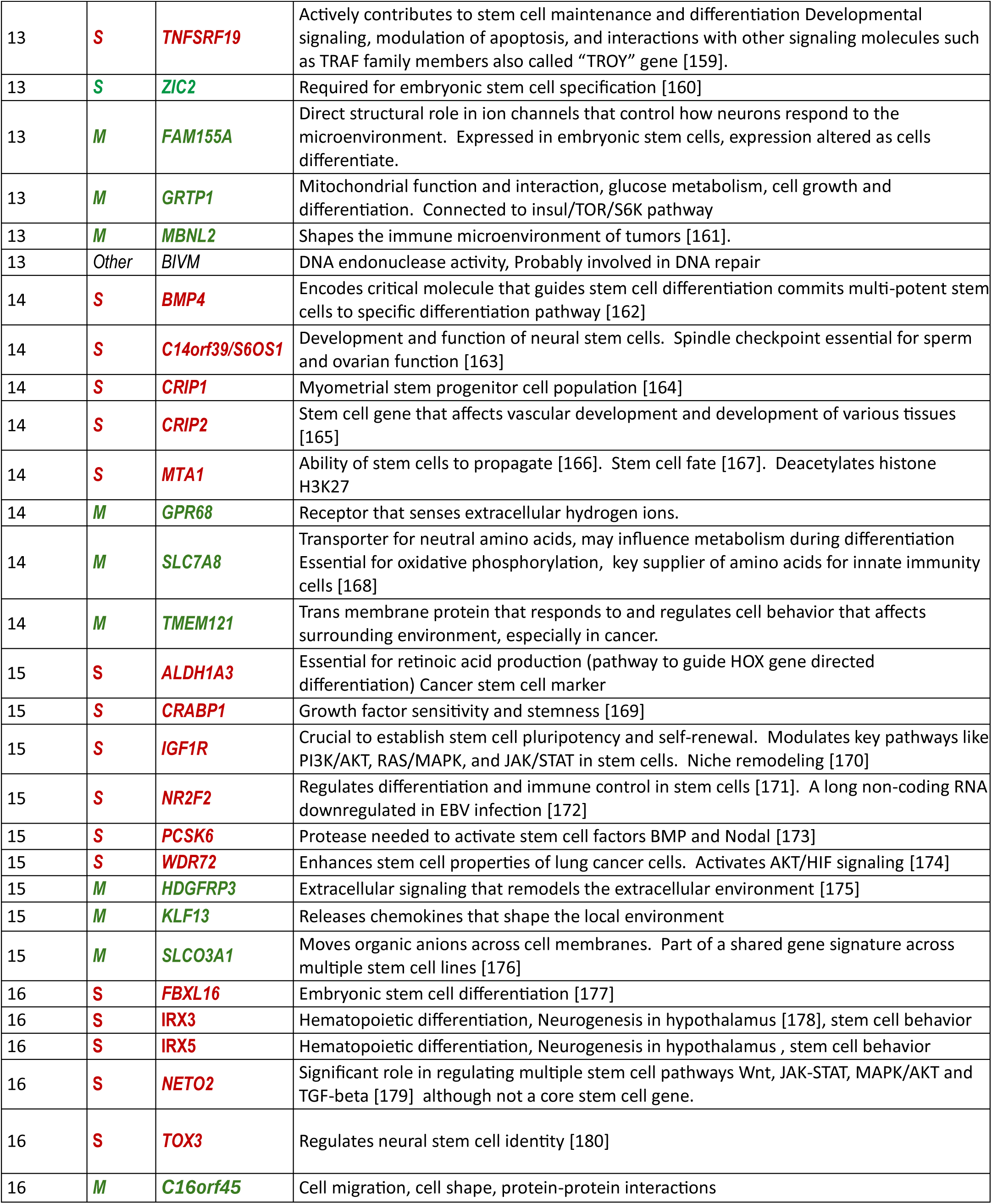
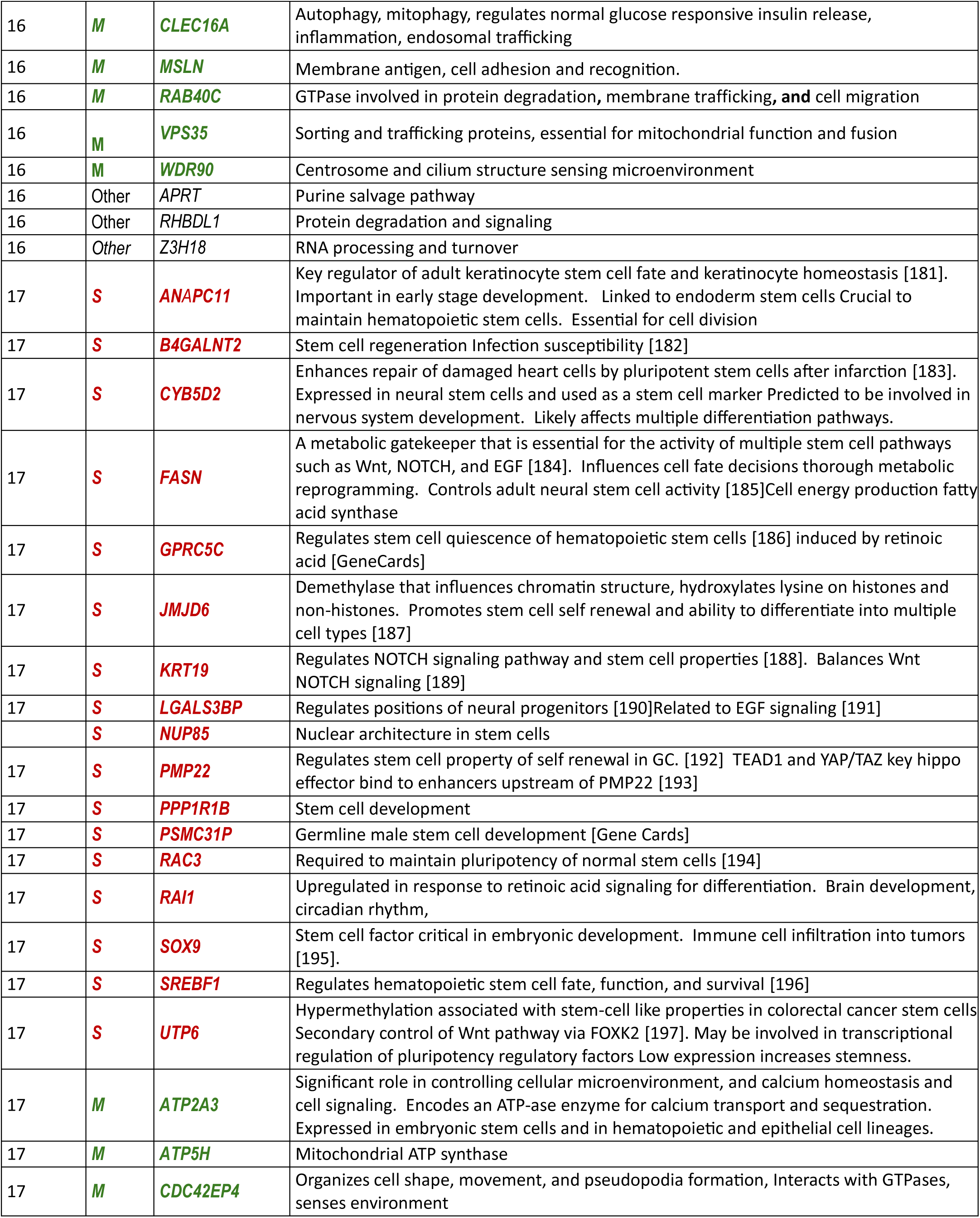
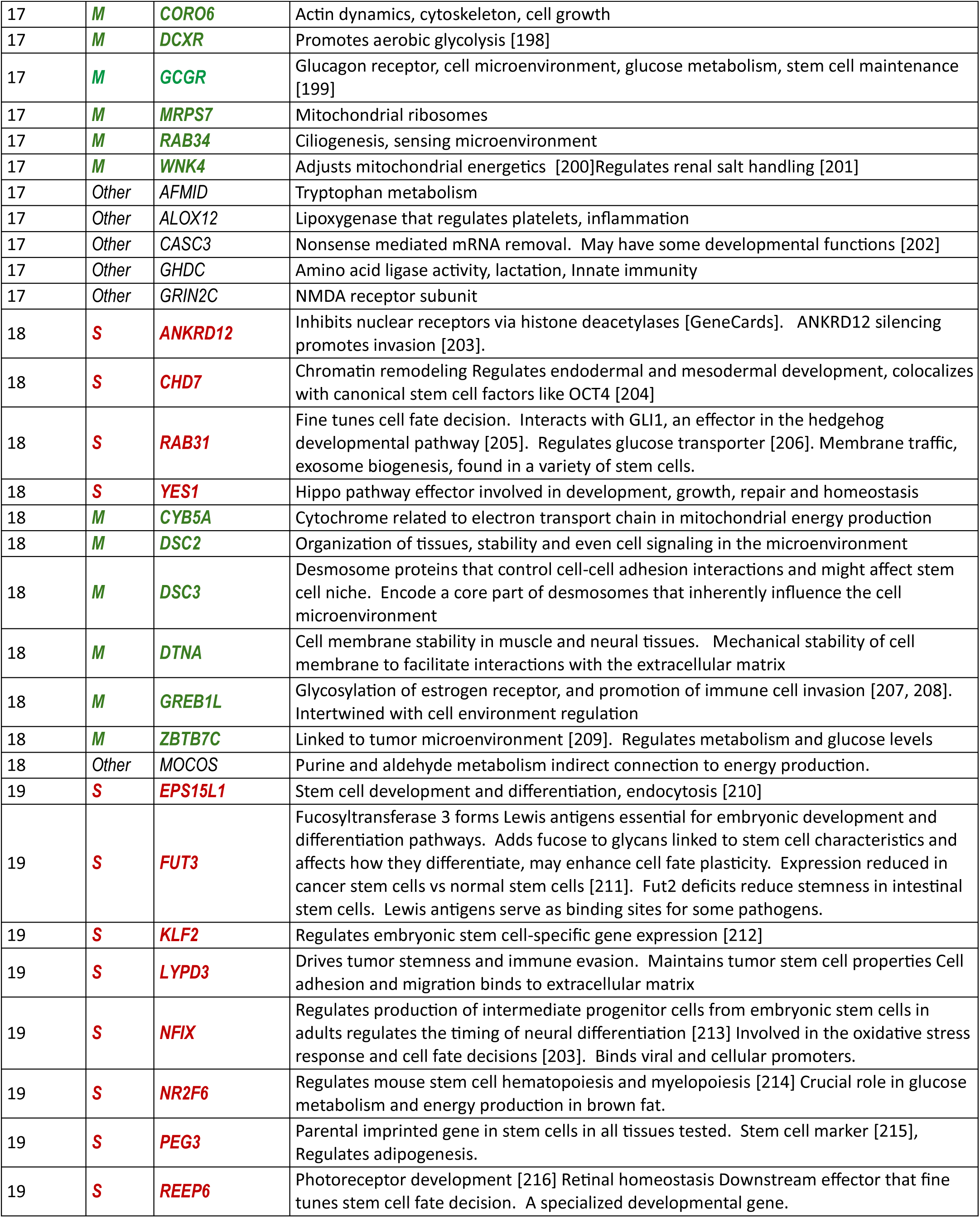
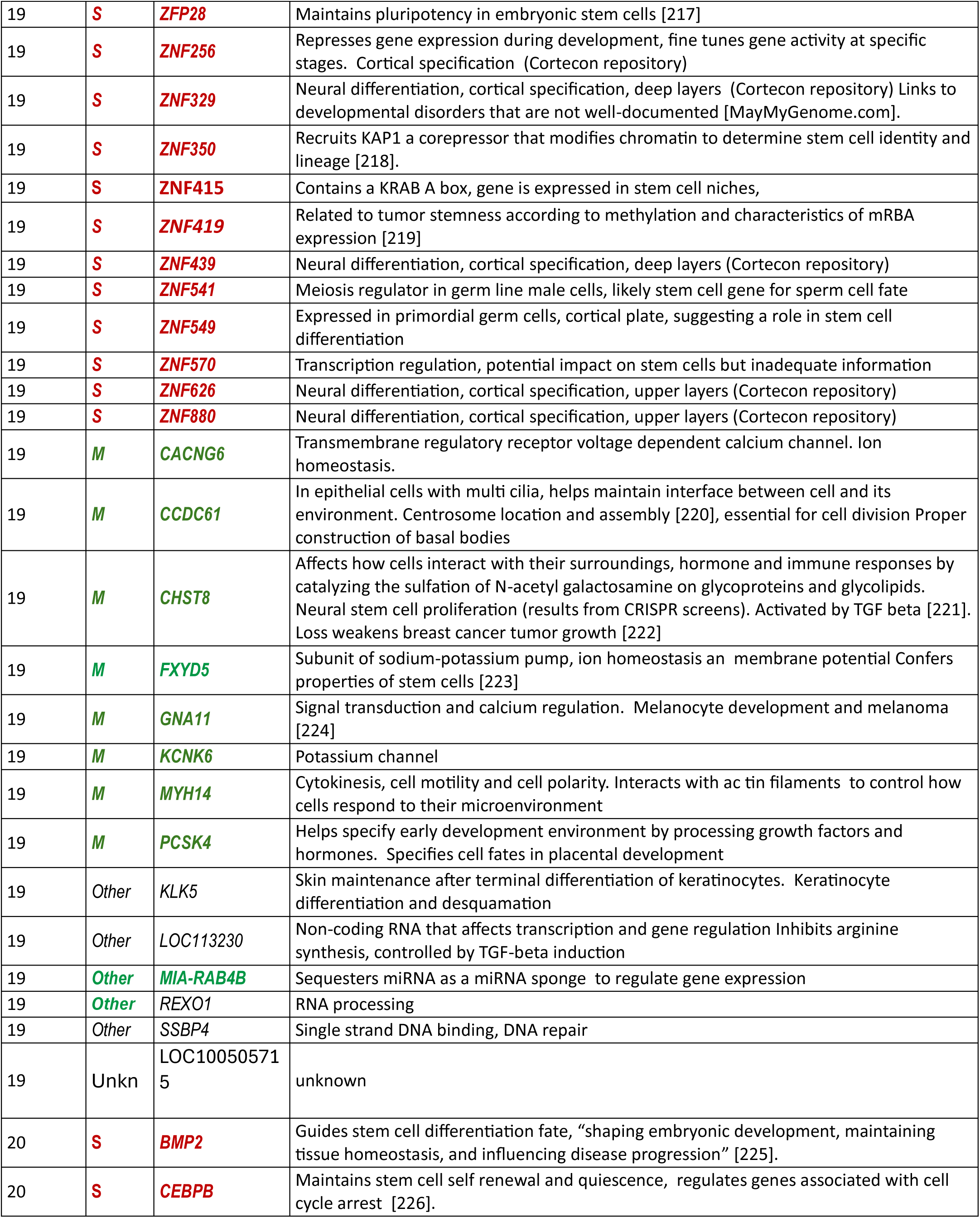
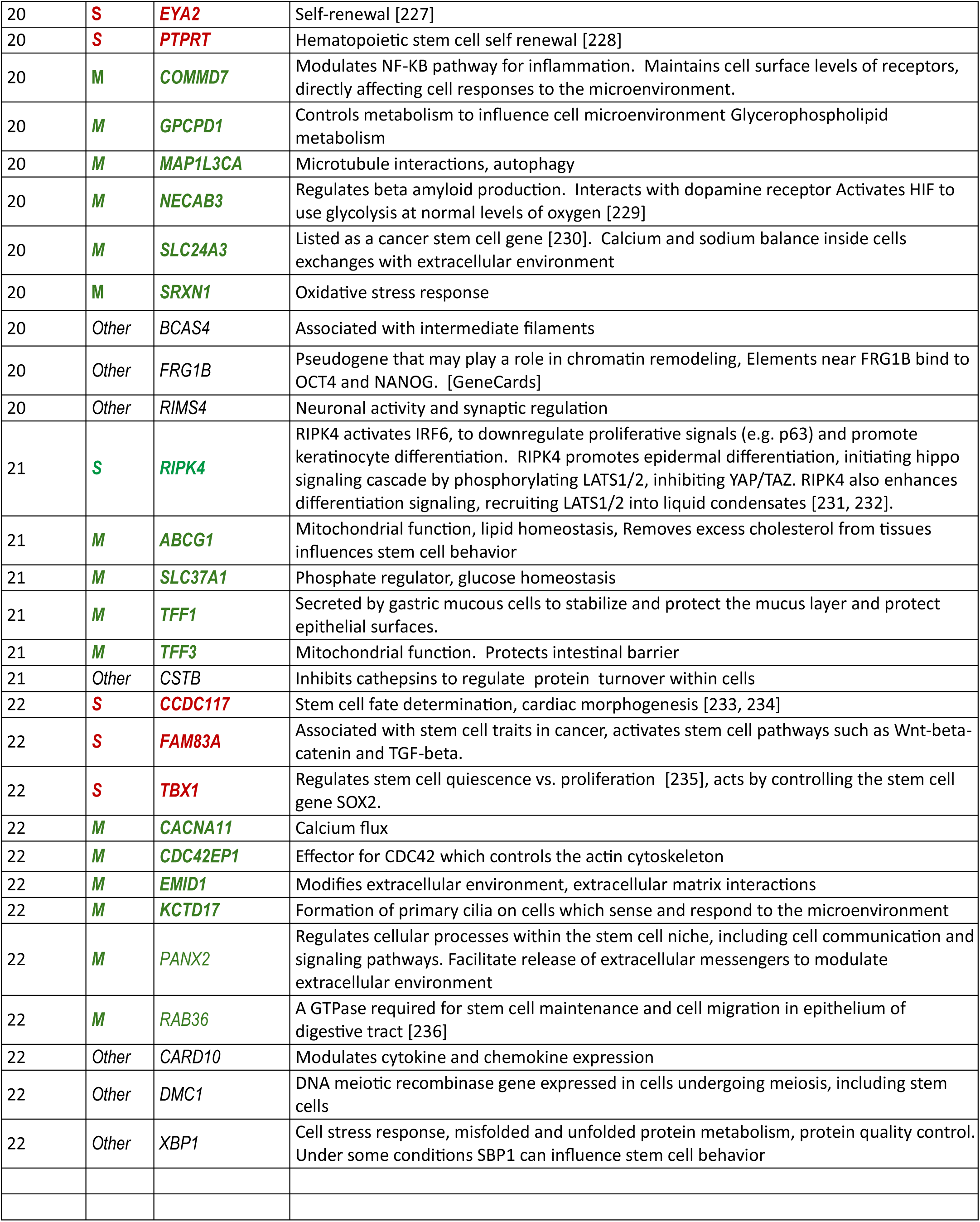
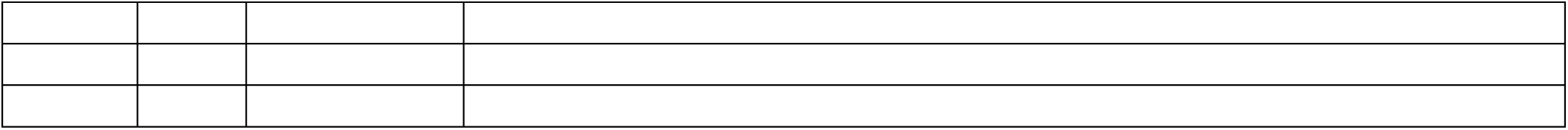
Matching abnormal methylation of gene controls in breast cancers and EBV cancers affects genes associated with stem cells and the microenvironment. S=Stem cell related, M=microenvironment, Other=not directly related to stem cells or microenvironment.

**Supplementary Table S2.** Functions of gene control sites methylated in non-malignant oral keratinocytes that are not methylated in the breast cancer cohort.

| **Chromosome** | **Gene** | **Class** | **Encoded Function** |
| --- | --- | --- | --- |
| 2 | *CYP1B1* | Microenvironment | Detoxification |
| 2 | *CYP27A1* | Other | Cholesterol metabolism |
| 2 | *EMX1* | Stem cell | Stem cell self renewal and proliferation |
| 2 | *GPR39* | Microenvironment | Zinc homeostasis |
| 2 | *MCEE* | Microenvironment | Metabolic enzyme essential for energy production in mitochondria |
| 4 | *CHRNA9* | Microenvironment | Ion channel receptor for acetyl choline expressed in stem cells |
| 5 | *EGFLAM* | Microenvironment | Extracellular matrix organization |
| 5 | *STC2* | Microenvironment | Calcium phosphate homeostasis |
| 5 | *ZNF354A* | Other | Transcription regulator cancer biomarker |
| 6 | *MB21D1* | Other | Antiviral defense, DNA damage sensor |
| 7 | *KCNH2* | Microenvironment | Voltage gated potassium channel |
| 8 | *HEY1* | Stem cell | Downstream target of Notch signaling pathway, regulates self renewal and differentiation |
| 8 | *NAT2* | Other | Drug metabolism and detoxification |
| 8 | *ZNF1* | Other | Transcription regulation |
| 9 | *GLDC* | Microenvironment | Glycine catabolism, redox balance |
| 9 | *SLC1A1* | Microenvironment | Glutamate transporter essential for neural stem cell self-renewal [1] |
| 9 | *TPRN* | Other | Inner ear function, cell structure and function |
| 10 | *GOT1* | Microenvironment | Glutamate level regulation, redox balance |
| 10 | *RNLS* | Microenvironment | Regulates cardiac function, blood pressure and immune microenvironment. |
| 10 | *UTF1* | Stem cell | Maintains pluripotency in embryonic stem cells [2] |
| 11 | *DAGLA* | Microenvironment | Affects hormone and immune microenvironment in the CNS |
| 11 | *MAP4K2* | Stem cell | Member of a pathway that affects cell fate of pluripotent stem cells |
| 11 | *MUC6* | Microenvironment | Forms gelatinous barrier to protect epithelial cells, alters cell signaling, modifies ECM and immunity |
| 14 | *GPX2* | Microenvironment | Redox balance and detoxification |
| 15 | *ANPEP* | Microenvironment | Endocytosis, amino acid and peptide transport. Used as a stem cell marker |
| 16 | *MT3* | Microenvironment | Detoxification, cell response to stress, zinc homeostasis. May be involved in deciding cell fate. |
| 16 | *POLR3K* | Other | Transcription, Subunit in RNA polymerase III. |
| 17 | *CLDN7* | Microenvironment | Mitochondrial tight junctions |
| 17 | *RAC3* | Stem cell | Required to maintain pluripotency of normal stem cells [3] |
| 17 | *RTN4RL1* | Microenvironment | Signaling cascades that impact actin cytoskeleton. May have stem cell like properties in nervous system but too little information available |
| 19 | *EPOR* | Stem cell | Stem cell differentiation especially in erythrocyte and neural progenitor cells [4]. |
| 19 | *GAMT* | Other | Creatine synthesis, muscle function, energy metabolism |
| 19 | *ZNF576* | Other | Transcription regulation |
| 22 | *A4GALT* | Microenvironment | Production of a glycolipid called globotriaosylceramide Membrane composition and receptor availability for cell interactions. |
| 22 | *MAPK8IP2* | Stem cell | Affects cell fate decisions in pluripotent neural progenitor cells, probably affects JNK signal pathway [5] |
| 22 | *RASD2* | Stem cell | Stem cell differentiation pathways |
| 22 | *SOX10* | Stem cell | Stem cell regulator and marker, mammary stem cells [6] |

